# An umbrella review of the evidence linking oral health and systemic health: from the prevalence to clinical and circulating markers

**DOI:** 10.1101/2022.04.11.22273715

**Authors:** João Botelho, Paulo Mascarenhas, João Viana, Luís Proença, Marco Orlandi, Yago Leira, Leandro Chambrone, José João Mendes, Vanessa Machado

**Author notes:** Corresponding Author (all below information can be published): João Botelho, Clinical Research Unit (CRU), Centro de Investigação Interdisciplinar Egas Moniz (CiiEM). Egas Moniz - Cooperativa de Ensino Superior, CRL, Address: Campus Universitário, Quinta da Granja, Monte de Caparica, 2829 - 511 Caparica, Almada, Portugal.; Fax: not available.

## Abstract

Oral conditions are highly prevalent worldwide. Recent studies have been supporting a potential bidirectional association of oral disorders with systemic noncommunicable diseases (NCDs). Robust evidence supports the greater prevalence of oral conditions in people suffering from NCDs limiting the ability of oral self-care. As for the relationship with other NCDs, the lines of evidence have increased exponentially but not always with the proper coherence. This umbrella review of meta-analyses appraises the strength and validity of the evidence for the association between oral health and systemic health. An extensive search included systematic reviews that have provided meta-analytic estimates on the association of an oral condition with a NCD. The overall strength of evidence was found to be unfavorable and with methodological inconsistencies. Twenty-two NCDs, four types of cancer and circulating levels of CRP were strongly associated with oral diseases. Among the significant NCDs are diabetes mellitus, cardiovascular diseases, depression, neurodegenerative conditions, rheumatic diseases, inflammatory bowel disease, gastric helicobacter pylori, stroke, obesity, diabetes mellitus or asthma. Most evidence is unlikely to change which indicates a relatively robust consistency of the available body of evidence.

## Introduction

Oral conditions are highly prevalent worldwide, a clear public health concern that disproportionately affects poorer and socially-disadvantaged populations [1–8]. Beyond its pronounced prevalence and burden, oral diseases have been proposed to have a bidirectional association with systemic health only suggested in recent years [9–11]. While evidence on this bidirectional link is robust on diseases that limit oral self-care (either physical or cognitive incapacity), the association of oral conditions with other chronic non-communicable illnesses has increased, still without the proper consistency.

The World Health Organization (WHO) approved, in 2021, a Resolution on oral health, urging key risk factors of oral diseases shared with other noncommunicable diseases (NCDs) [12]. Instead of the traditional curative approach, WHO caveats the importance of prevention encompassing oral health in universal health coverage programs. With estimates of more than 3.5 billion people suffering from oral diseases, the associated burden is likely to remain or increase, particularly during the current global pandemic, with successive lockdowns that have led to limitation of access to oral health care. Analyzing consequently the degree of evidence of these diseases with systemic pathologies becomes highly relevant to inform and influence public health and health policy makers. For this reason, we performed an umbrella review to overlook the robustness of the meta-analytic estimates linking oral and systemic conditions. We additionally explored whether future research will likely transform the inferences from existing significant meta-analyses.

Herein, we demonstrate that twenty-two NCDs, four types of cancer and circulating levels of CRP were strongly associated with oral conditions. Most evidence is unlikely to change in the future, with a few exceptions, which indicates relatively robust consistency of evidence provided.

## Results

### Selection and characteristics of the included meta-analyses

Our search retrieved a total of 13,342 entries (Figure 1). After removing duplicates (n=7,884), a total of 5,458 records were screened for title and abstracts against the eligibility criteria. After judging the full-paper of 591 records, 378 studies were excluded (the list of excluded studies with justification for exclusion is detailed Supplementary Data 1). Excellent inter-examiner reliability was confirmed at the full-text screening (kappa score = 0.91, 95% CI: 0.88; 0.93). A final sample of 213 meta-analyses were included for further appraisal (Supplementary Data 2).

**Figure 1.**
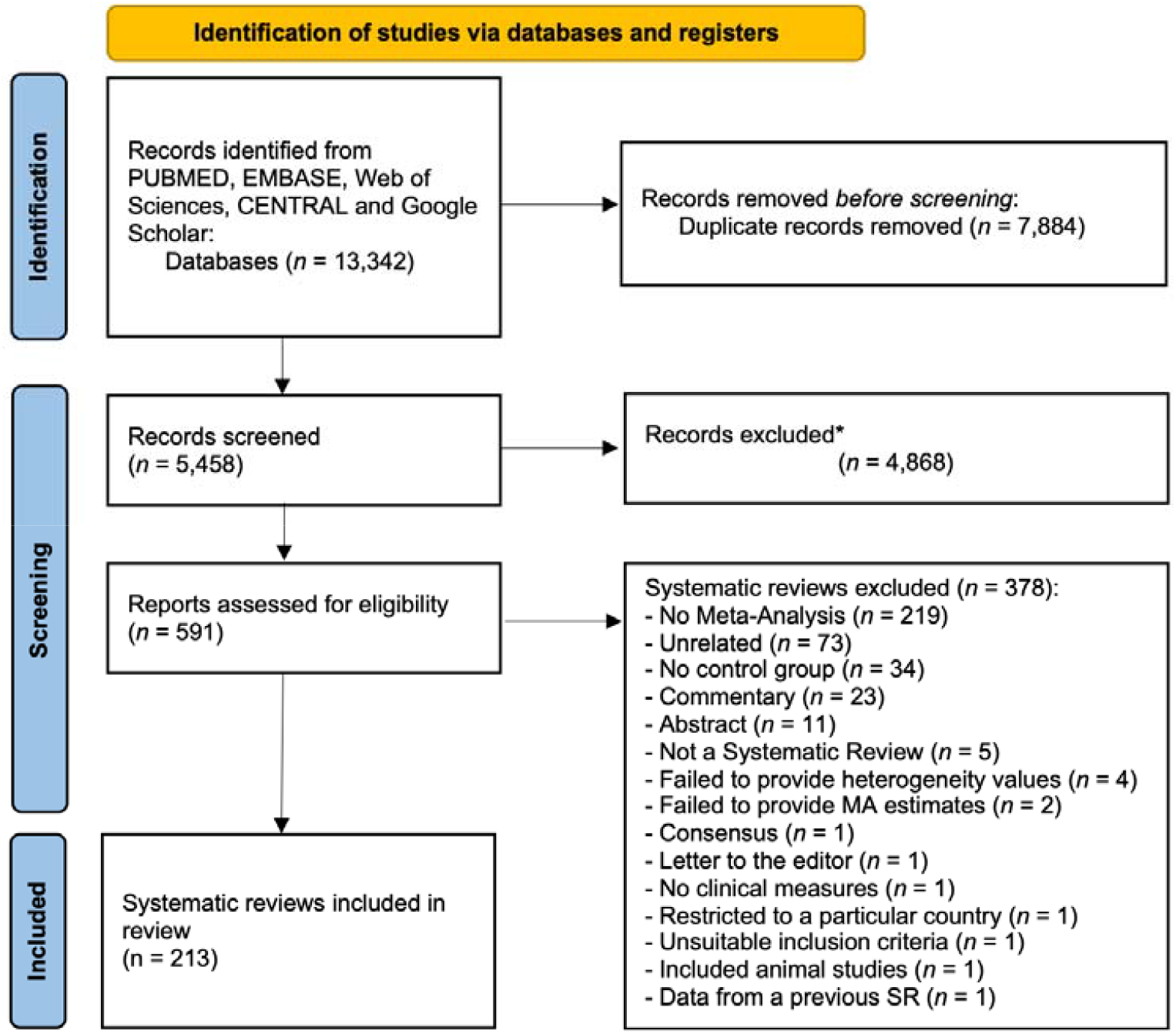
PRISMA flowchart showing the exclusion and inclusion process of the literature review.

Most meta-analyses were conducted in Brazil (n=43), China (n=42), United Kingdom (n=18), USA (n=17), Spain (n=14) and Italy (n=10), still we observed studies from a substantial number of countries (Supplementary Data 3). The majority followed Preferred Reporting Items for Systematic Reviews and Meta-Analysis (PRISMA) (n=145, 75.1%), Meta-analyses Of Observational Studies in Epidemiology (MOOSE) (n=17, 8.8%), or both two or three guidelines at the same time (PRISMA, MOOSE and/or Cochrane) (n=15, 7.0%). Yet, fourteen did not report following a guideline for systematic reviews (n=14, 7.3%). While for risk of bias (methodological quality), Newcastle-Ottawa Scale (NOS) (n=95, 44.6%), Cochrane tools (n=27, 12.7%) or Joanna Briggs Institute (JBI) tools (n=10, 4.7%) were the most used instruments.

Overall, 634 meta-analytic comparisons were included. Half meta-analyses (n=315; 50.0%) used a continuous exposure contrast, whereas the other half used a binary analysis (Supplementary Data 2). Most meta-analyses used oral conditions and/treatments as exposure (n = 377, 59.5%), while the remaining used them as outcome. The summary descriptive characteristics of the included meta-analyses by oral condition is presented in Table 1.

### Summary effects and heterogeneity between studies

Of the 634 meta-analytic comparisons, 430 (67.8%) were nominally significant (p<0.05), with only 8.5% of strong meta-analytical evidence, while 19.6% (n=124) and 6.2% (n=39) were of highly suggestive and suggestive evidence, respectively (Table 1). Of the stricter P value threshold, 276 (43.5%) and 82 (12.9%) meta-analyses had significance at 10^−6^ and 10^−3^, respectively. Approximately 62% (n=395) of the included meta-analyses had high heterogeneity (I^2^>50%), with almost one fourth of them presenting low heterogeneity (I^2^≤25%).

### Grading of the evidence from oral conditions

Fifty-four meta-analyses (8.5%) were categorized as of strong evidence (Figures 2-5), thirty-five with oral diseases as exposure towards a particular systemic disease and nineteen with oral conditions as outcome. Considering oral conditions as an exposure of a particular systemic condition, the following associations were found: dental caries with iron deficiency (n=2); tooth loss with cognitive impairment (n=2), with dementia (n=2) and with lung cancer (n=1); edentulism with pancreatic cancer (n=1); endodontic infection with serum C-reactive protein (CRP). In addition, periodontal disease presented strong association with higher risk towards: cancer (n=5); cardiovascular disease (CVD) (n=4); diabetes mellitus (n=3); adverse pregnancy outcomes (APOs) (n=2); lower longevity (n=2); neurodegeneration (cognitive impairment and dementia) (n=2); polycystic ovarian syndrome (PCOS) (n=1); psoriasis (n=1); altered levels of mean corpuscular hemoglobin (MCH) (n=1); and, with serum CRP (n=1). Comparing with healthy counterparts, the following associations were found considering oral condition as outcome: mental disorders with dental caries (n=2) and tooth loss (n=1); conditions of special needs with dental trauma (n=1); diabetes mellitus with denture stomatitis (n=1) and periodontal disease (n=1); CVD with higher average tooth loss (n=1); asthma with mouth breathing (n=1) and higher average of gingival bleeding (n=1); and, obese patients with edentulism (n=1). As well, this risk towards periodontal disease was found associated with: lower physical activity (n=1); rheumatoid arthritis (n=1); PCOS (n=1); nonalcoholic fatty liver disease (n=1); medication-related osteonecrosis of the jaw (MRONJ) (n=1); ankylosing spondylitis (n=1); inflammatory bowel disease (n=2); and, gastric *helicobacter pylori* (n=1).

**Figure 2.**
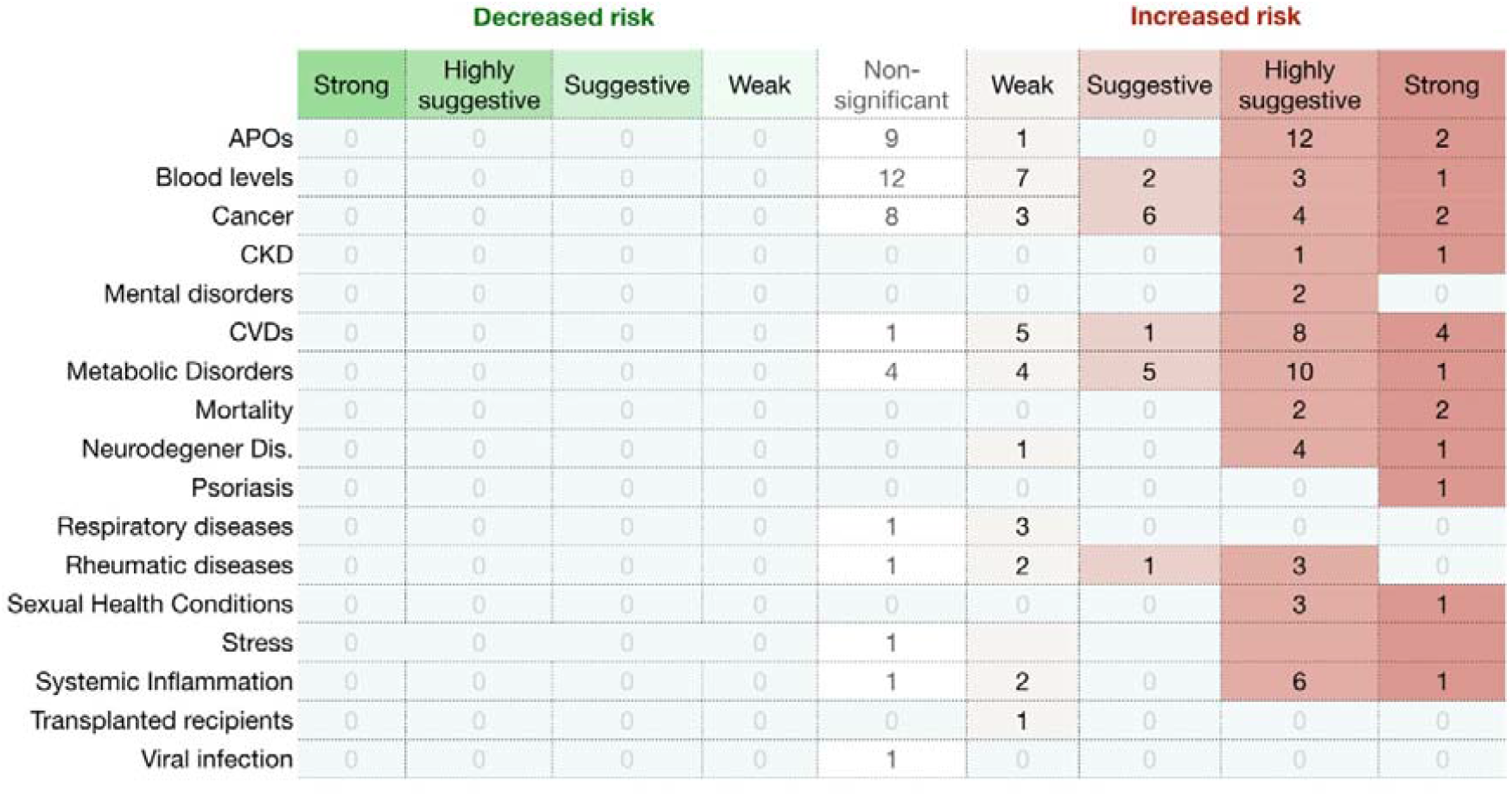
Evidence grading diagram on periodontal diseases as an outcome of a respective systemic NCD. The right side displays associations that increase the risk for the respective systemic NCD (in red), whereas the left side shows associations that reduce the risk (in green). APO adverse pregnancy outcomes, CVDs cardiovascular diseases, CKD chronic kidney disease, Dis. disease.

**Figure 3.**
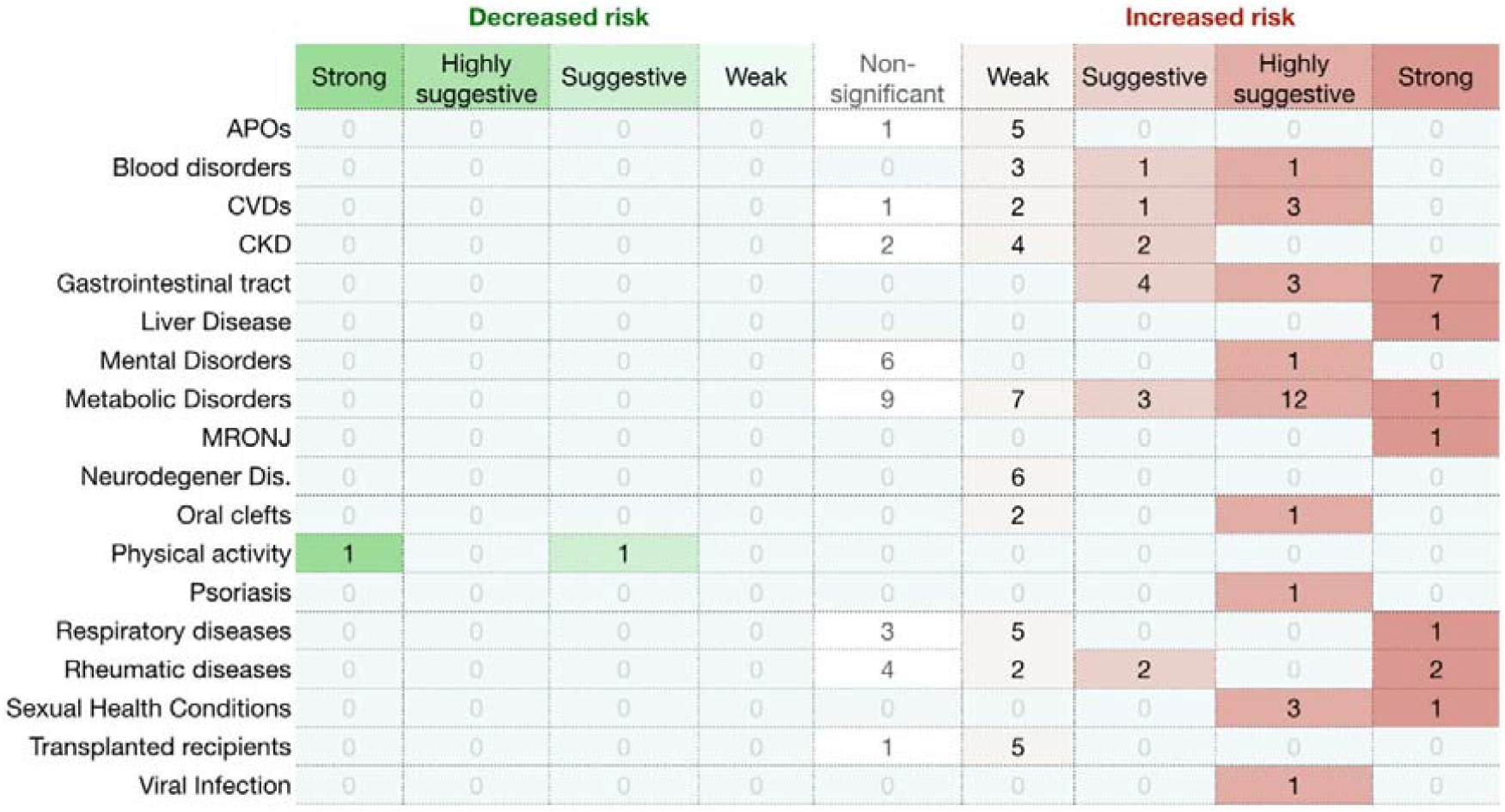
Evidence grading diagram on periodontal diseases as an exposure to a systemic NCD. The right side displays associations that increase the risk for the respective systemic NCD (in red), whereas the left side shows associations that reduce the risk (in green). APO adverse pregnancy outcomes, CVDs cardiovascular diseases, CKD chronic kidney disease, Dis. disease.

**Figure 4.**
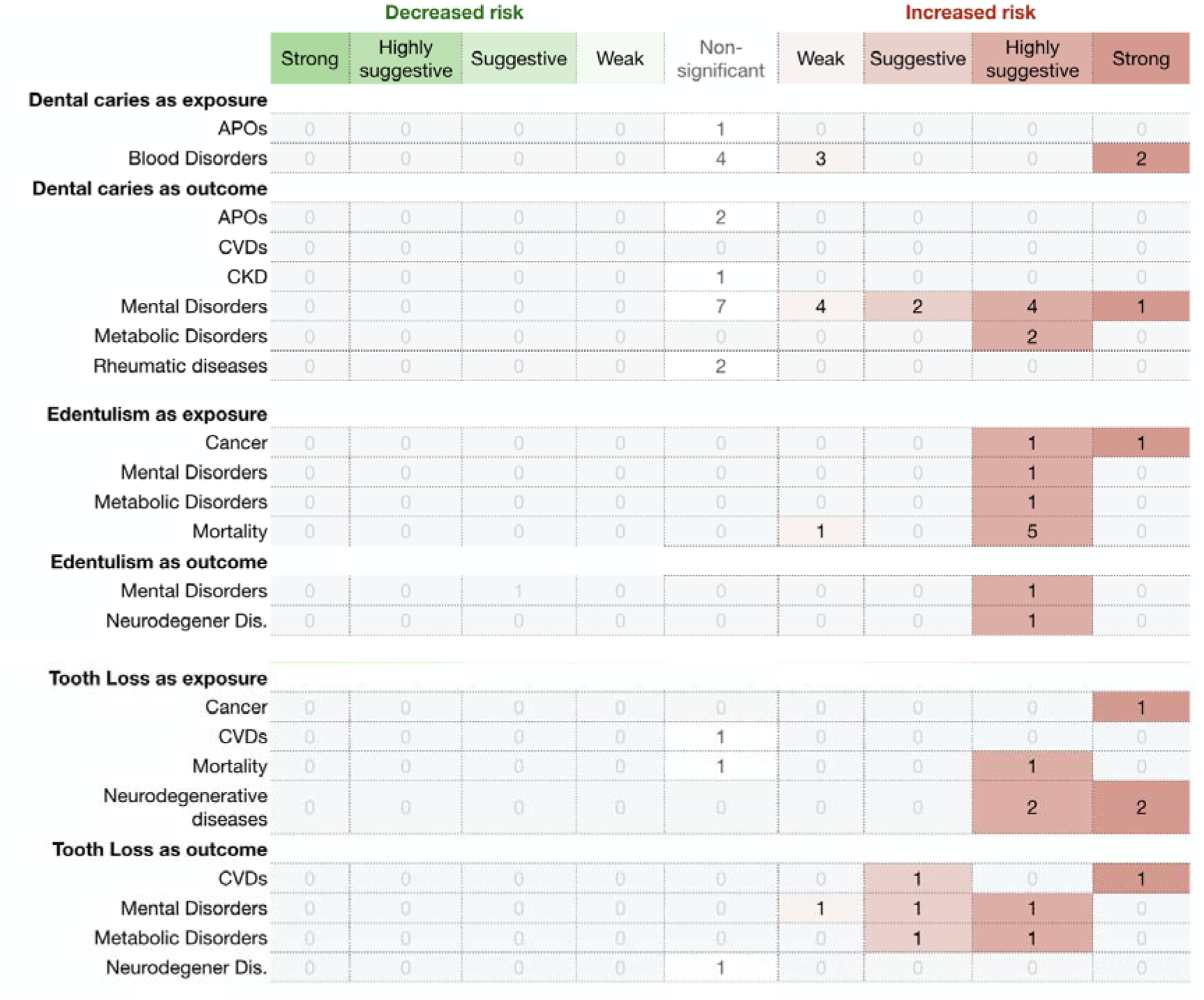
Evidence grading diagram on dental caries, edentulism and tooth loss (both as an exposure and outcomes) of a systemic NCD. The right side displays associations that increase the risk for the respective systemic NCD (in red), whereas the left side shows associations that reduce the risk (in green). APO adverse pregnancy outcomes, CVDs cardiovascular diseases, CKD chronic kidney disease, Dis. disease.

**Figure 5.**
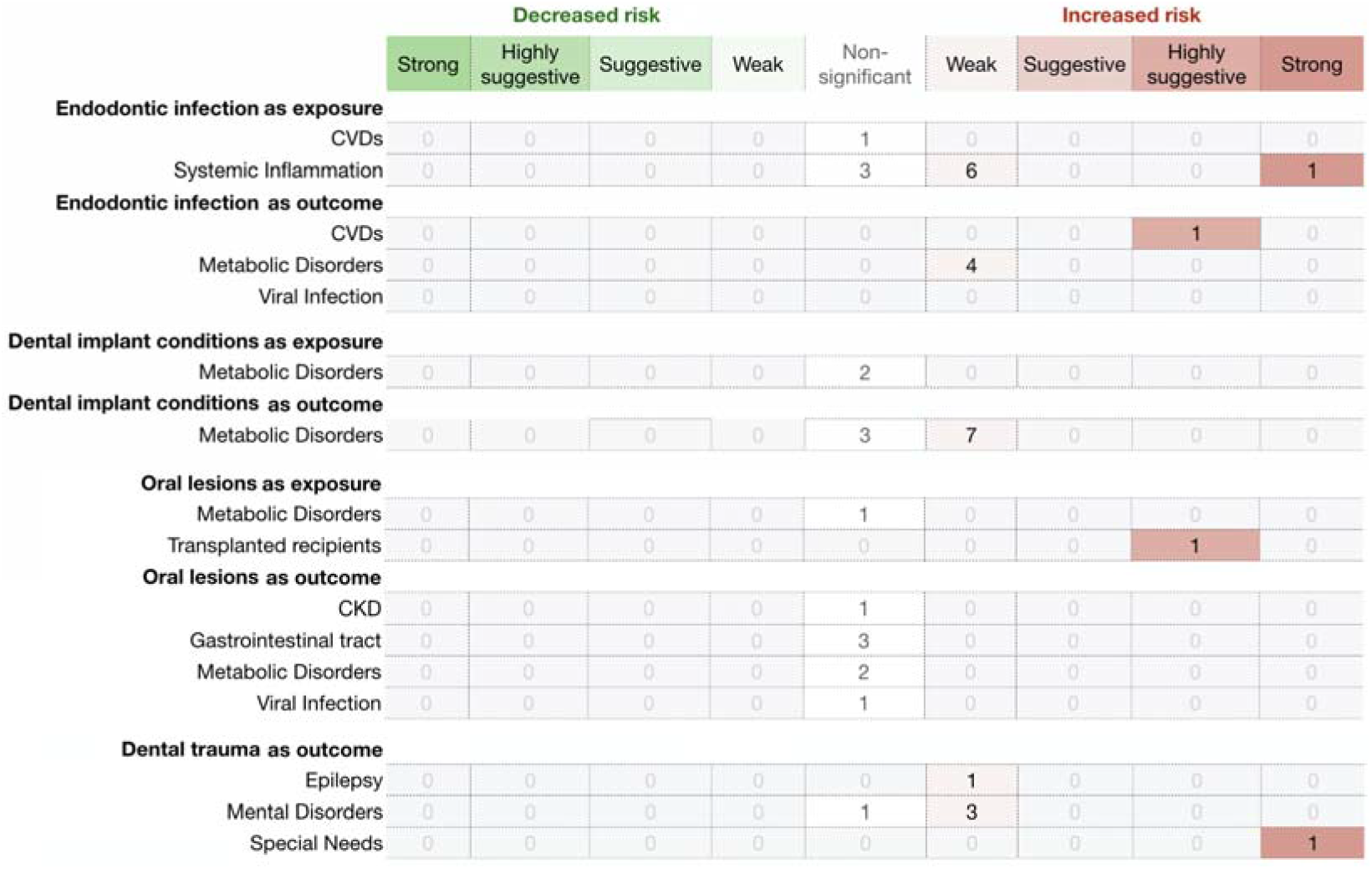
Evidence grading diagram on endodontic infection, dental implant conditions, oral lesions and dental (both as an exposure and outcomes) of a systemic NCD. The right side displays associations that increase the risk for the respective systemic NCD (in red), whereas the left side shows associations that reduce the risk (in green). APO adverse pregnancy outcomes, CVDs cardiovascular diseases, CKD chronic kidney disease, Dis. disease.

### Grading of the evidence from the impact of oral treatments

A total of 149 meta-analyses (23.5%) explored the impact of an oral treatment (either periodontal, endodontic or dental in general) on systemic diseases and/or makers (Table 2). Only two had strong meta-analytical evidence: periodontal treatment on systemic inflammation (n=1); and endodontic treatment on CVDs (n=1) (Figure 8 displays the map of evidence on these associations).

**Figure 8.**
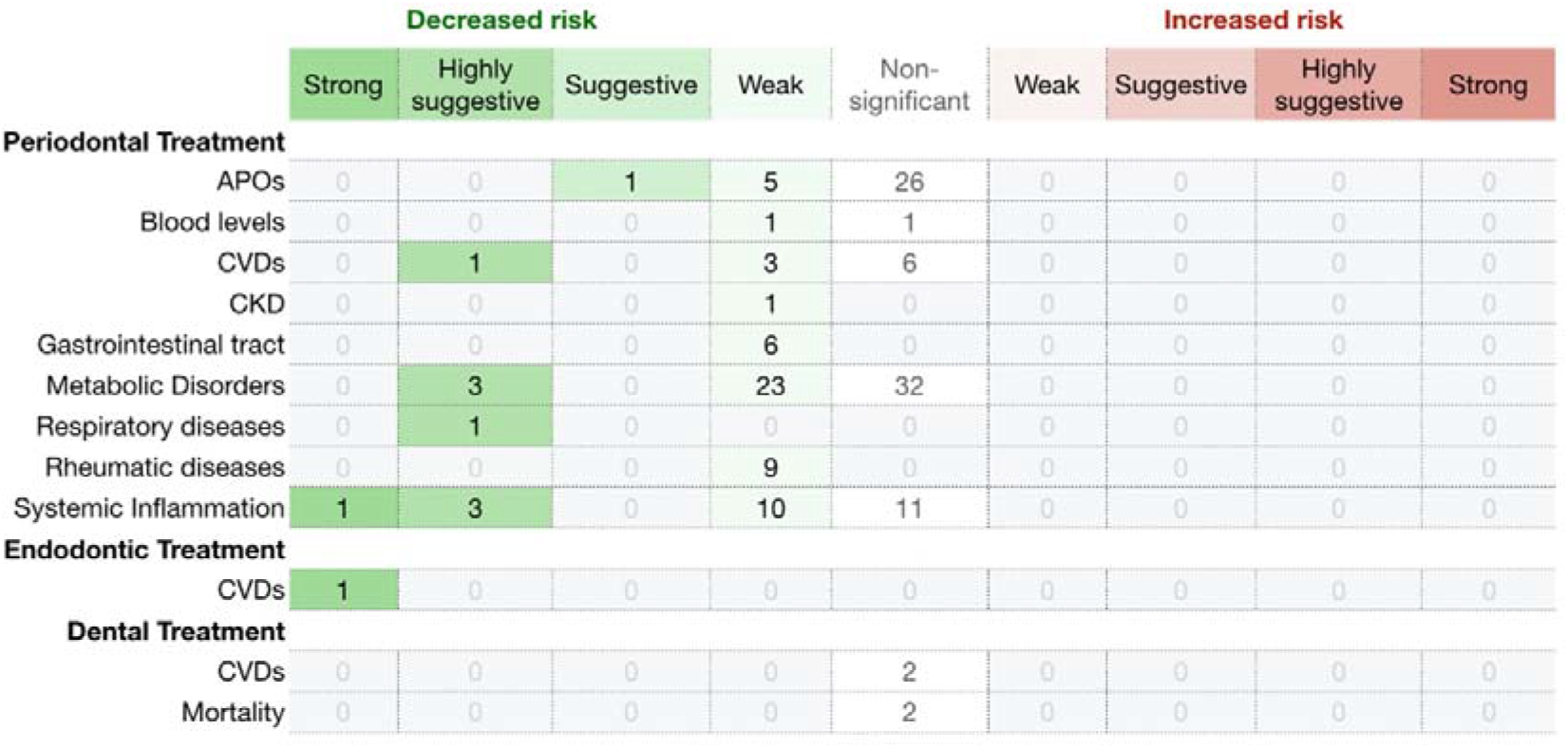
Diagram showing results from the umbrella review grading the evidence on diet and cancer risk. The right side displays associations that increase the risk for the respective systemic NCD (in red), whereas the left side shows associations that reduce the risk (in green). APO adverse pregnancy outcomes, CVDs cardiovascular diseases, CKD chronic kidney disease.

Furthermore, periodontal treatment presented highly suggestive evidence of impacting CVDs (n=1), metabolic disorders (n=3), respiratory diseases (n=1) and systemic inflammation (n=3), as well as suggestive evidence on APOs (n=1). In addition, weak evidence was found on the effect of periodontal therapy on APOs (n=5), blood levels (n=1), CVDs (n=3), chronic kidney disease (CKD) (n=1), gastrointestinal traits (n=6), metabolic disorders (n=23), rheumatic diseases (n=9) and systemic inflammation (n=10).

### Methodological quality assessment

Good inter-examiner reliability at the A Measurement Tool to Assess Systematic Reviews 2 (AMSTAR 2) screening was recorded (kappa score = 0.83; 95% confidence interval (CI): 0.79-0.86). Only fifteen meta-analyses were conducted with high methodological quality (7.0%) and eight with moderate (3.8%), according to this appraisal tool (Supplementary Data 4). The majority presented low (n=42, 19.7%) to critically low methodological quality (n=148, 69.5%). The included meta-analyses predominantly failed to report on the funding sources for the studies included in the review (n=197, 92.5%), to assess the potential impact of risk of bias in the meta-analysis (n=158, 74.2%), to list the excluded studies with the respective justification (n=147, 69.0%) and to account risk of bias in the interpretation and discussion of the results (n=132, 62.0%). At a lower proportion, but also seriously, a comprehensive literature search strategy was also lacking in 33.3% of the included studies (n=71), while the definition of the review methods *a priori* was not accounted in 20.2% (n=43). The selection (n=36, 16.9%) and data extraction (66, 31.0) in duplicates was absent as well. Publication bias was not performed in 26.8% (n=43) of meta-analyses, even if there were prerequisites to do so.

### Number of additional studies needed to change current meta-analytic evidence

Among the 219 meta-analyses that achieved from suggestive to strong evidence, the median fail-safe number (FSN) was 47 (range: 1 to 87,973). For each level of evidence, the median FSN was 14 (range: 1 to 80) for suggestive, 61 (range: 2 to 87,973) for highly suggestive and 59 (range: 5 to 1,618) for strong evidence (Supplementary Data 5). The FSN was larger than the number of studies included in in 98.2% of the meta-analyses (n=215) for these evidence categories, meaning the statistically significance of the summary estimates is highly unlikely to change if a number of studies are further added in the future. Regarding the 210 weak evidence meta-analyses, the median FSN was 11 (range: 0 to 1,501), with the FSN being smaller than the number of included studies in the existing meta-analyses in 50 comparisons (23.8%).

## Discussion

### Summary of main results

The present umbrella review assessed a total of 213 meta-analyses with a total sample of 634 comparisons. Fifty-five associations were considered of strong evidence, supported by highly significant results and absence of bias. The available evidence allowed to group strong evidence in three main categories: i) people suffering from NCDs at a higher risk towards oral conditions; ii) the exposure of an oral condition increasing the risk towards a systemic NCD; iii) the systemic effect of oral interventions. Thirteen systemic NCDs were associated with a higher risk of having an oral condition: depression (with dental caries and tooth loss), several mental disorders (with dental caries), rheumatoid arthritis (with periodontal disease), anquilosis espondilitis (with periodontal disease), inflammatory bowel disease (with periodontal disease), nonalcoholic fatty liver disease (NAFLD) (with periodontal disease), PCOS (with periodontal disease), MRONJ (with periodontal disease), gastric helicobacter pylori (with periodontal disease), stroke (with tooth loss), obesity (with edentulism), diabetes mellitus (with denture stomatitis), asthma (with periodontal disease and mouth breathing) and various conditions of special needs (with dental trauma). Physically active people were associated with a lower likelihood towards periodontitis. Regarding the role of oral conditions on systemic health, most associations pertained to periodontal diseases with APOs, diabetes mellitus, gestational diabetes mellitus, CKD, CVD, PCOS, dementia, psoriasis, cancer (pancreas, prostate, lung, head and neck), cognitive decline and dementia. Other associations, particularly tooth loss and edentulism (with pancreatic and lung cancer, cognitive decline and dementia), and dental caries with iron deficiency. In addition, both periodontal and endodontic infection were confirmed as sources of systemic inflammation, as well periodontal disease was linked to changes in MCH. Regarding intervention evidence, periodontal and endodontic procedures were strongly associated with improvements in circulating levels of CRP and lower risk towards CVD, respectively. All in all, a total of twenty-two NCDs, 4 types of cancer and circulating markers of inflammation were strongly associated with oral conditions.

Periodontitis account for the majority of the associations with systemic NCDs and markers, followed by tooth loss and edentulism, in some cases indirect measures of periodontitis. The rise of Periodontal Medicine as an independent field in periodontal research can account for the relatively significant portion of such studies on periodontology [13]. While the mechanisms within this association (see subsection ‘***The mechanisms within the oral-systemic intersection’***) have been progressively studied and understood, this bulk of knowledge has expanded to other areas, such as Peri-implantology and Endodontics. Primarily, the pathophysiology of periodontitis seems to be similar in peri-implantitis and apical periodontitis for endodontic reasons, so it will not be surprising, in the near future, for studies on the intersection of these pathologies with systemic diseases to draw a parallel with periodontitis. With respect to tooth loss and edentulism, both are clinical endpoints of mainly periodontal diseases, and to a minor extent of dental caries.

Through methodological analysis and meta-analytic evidence, these results may have some degree of impact by biases. Only 10.8% of meta-analyses (n=23) were conducted with high/moderate quality according to AMSTAR 2. While some of the observed issues may have residual impact on the meta-analyses (such as, reporting funding from the included studies, accounting risk of bias on interpretation and discussion of the results, and defining the protocol *a priori*), others may have adverse effects to the consistency of the results (lack of a comprehensive search, absence of duplicate data search and extraction, and list of excluded studies with respective reason). Furthermore, from the statistical point-of-view, the included systematic reviews had on average relatively few studies (median=9) and few participants included (median=1,113). Nevertheless, more than 55% of the included comparisons reported statistically significant results, with a substantial number, about 38% presenting a lower *P* value threshold (*P*<10^−6^, n=239). Around 47% (n=297) showed high heterogeneity (*I*^2^>50%), and only 32.7% had low heterogeneity (*I*^2^≤50%). Through FSN metrics, we concluded that most evidence is unlikely to change, with a few exceptions, which indicates relatively robust consistency of evidence provided. For this reason, future research in the oral-systemic health intersection shall seek to strengthen compliance with established guidelines. Additionally, efforts should be made to increment the number of intervention trials to establish more conclusive inferences.

These results convey a sufficient knowledge on hypothetical observational associations that shall not be ignored, although there is still insufficient understanding on the conceivable causal role of oral conditions on systemic diseases, particularly, and vice-versa. In this regard, public health data has shown that preventative actions are effective both in preventing oral diseases as well as increasing quality of life [14]. Still, in line with these results, research on intervention on oral diseases and their systemic impact is still unfavorable and is of paramount importance to be included in the international research agenda. The access to oral care and inequity in oral health and disease worldwide, poorly addressed in the included systematic reviews, also requires attention. While most of this evidence derives from research from developed countries, contributions from developing countries are still unrepresentative. While this is slowly being normalized, with the substantial increase in research by developing countries, these results should reflect the importance of access to oral health care, preventive and curative measures, and the potential impact on systemic health it may have. In parallel, many of the systemic NCDs that showed strong evidence of association are both highly prevalent and with a disproportional incidence rate and an exacerbated impact in underdeveloped nations [15,16].

### The mechanisms within the oral-systemic intersection

Oral conditions might have an impact on systemic health via multiple pathways [9]. The oral microbiome, its byproducts and their interaction with the host immune system have been identified as the major players in this causal association. The largest body of evidence currently available focus on periodontitis as bacteremia and systemic inflammation represent plausible mechanisms of causality for the contribution of periodontitis to the pathogenesis of multiple diseases [9]. Chronic inflammation coupled with a richly vascularized periodontium leads to ulceration of the oral epithelial barrier, and consequently, greater access for pathogenic microbes and their products to the bloodstream [17]. Generally, the chronic systemic distribution of oral bacteria-derived products converges at the point of an altered state of immunity, achieved either through subversion of host defences, or prolonged and/or enhanced inflammatory responses. Low-grade systemic inflammation has been linked to the development of a wide spectrum of NCDs such as cardiometabolic, neurodegenerative, rheumatic and neoplastic conditions [18]. The increase in acute phase reactants and inflammatory cytokines could trigger the activation of immune cells such as circulating monocytes with a subsequent vascular inflammation [19]. Furthermore, chronic inflammation can cause alterations in lipid and lipoprotein metabolism leading to a proatherogenic lipoproteins setting [20]. In addition, systemic inflammation is associated to glucose intolerance and insulin resistance contributing to increase risk of type 2 diabetes mellitus and other metabolic disorders [21]. The action of oral bacteria such as *Porphyromonas Gingivalis (P. Gingivalis), Aggregatibacter Actinomycetemcomitans (A. Actinomycetemcomitans)* and *Fusobacterium Nucleatum (F. Nucleatum)* has been linked to the development of rheumatic conditions and certain types of cancer. *P. Gingivalis* and *A. actinomycetemcomitans* have shown the ability to trigger the production of anti-citrullinated protein antibodies (ACPAs) [22–24], a classic feature detected in rheumatoid arthritis. *F. Nucleatum* in *in vitro* and animal models can stimulate growth and migration of colorectal cancer (CRC) cells [25–28] and accelerates tumor growth and metastatic progression in breast cancer [29]. Furthermore, immune cells, specifically lymphocytes and myeloid cells, can be primed during the interaction with inflamed oral sites and have a detrimental effect at distant mucosal sites [30]. The effect of the treatment of periodontitis on several markers of systemic inflammation and disease activity, surrogate markers of cardiovascular health and metabolic control has corroborated the hypothesis of a causal association with NCDs [31–33].

### Strengths and Limitations

The protocol of the present umbrella review was developed *a priori* and registered in PROSPERO contributing to the robustness of its analyses and results, transparency and mitigation of errors. We did not limit this review to assessing the methodological quality but also focused on the degree of evidence of meta-analytic estimates based on the strategy defined by Papadimitriou and colleagues [34]. As argued, despite the existence of other methods for rating evidence quality, such as GRADE [35], this approach focuses on objective and tangible criteria. Furthermore, methodological quality via the AMSTAR2 allowed a comprehensive view from both randomized and non-randomized studies [36], and goes beyond the statistical sphere of meta-analyses. As well, the used of FSN allowed to learn that 98.4% of the significant evidence (from suggestive to strong) is unlikely to change even in the possibility of future research, thus allowing robust conclusions to be implemented in public health agenda and policymakers. Nonetheless, these indications aim to foster the interest in fill in the existing science and knowledge gaps, and that are still many given the relative youth of the intersection of oral health with health in general, and they do not intend to limit research in associations that achieved strong statistical association. In addition, this notion of completed science is unreal, as there are situations of strong associations with a short bulk of research carried out and highly suggestive or suggestive associations with a remarkable volume of research.

However, this review present important shortcomings worth discussing. The current umbrella review is heavily based on meta-analyses of observational studies and with a low percentage of randomized trials. Hence, the overall view of these results relies more on non-inferential evidence than causal definitive assumptions. Still, we highlight some of the observational studies included have a notable number of participants, and some, although scarce, have a prospective design and come from insurance databases with a population-level data source, such as the Taiwan’s National Health Insurance Research Database [37]. This sort of databases represents an opportunity to progress big data analysis by combining multiple groups of variables (physical, laboratorial, clinical or sociodemographic, among others). Nonetheless, the validity of the diagnosis codification codes (mostly based on international classification of disease 9 clinical modifications [ICD-9-CM] and ICD-10) are still controversial, because while the reliability of ICD-9-CM is somehow well established, ICD-10 codes are yet unvalidated [37].

Regarding the observational nature of the majority of studies included in the meta-analyses, there is a large variability in the measurement or diagnosis in commonly seen oral conditions, such as dental caries or periodontal disease. This heterogeneity may constitute a source of bias and may lead to under- or overestimated meta-analytical results. On the one hand, dental caries was mainly reported through the Decayed, Missing, and Filled Teeth (DMFT) index, the most commonly used epidemiological index for assessing dental caries [38]. While this index provides a complete view of the past history of caries, it overestimates caries experience by attributing the cause of all missing teeth to caries. In this case, assigning the maximum or minimum value is likely to over- or underestimate inequalities, respectively, and ignoring the ‘M’ component would omit possibly a major inequality component of this index [38]. On the other hand, the periodontal clinical measures, particularly clinical attachment loss (CAL) and periodontal probing depth (PPD), are often provided as a global sum of the patient’s mouth over thresholds of pathological PPD or CAL within the whole mouth [39]. Likewise, the variability of case definitions of periodontitis and gingivitis is also a possible source of bias. Recently a joint consensus case definition for periodontitis was proposed [40], and the variation from previous case definitions was attested using graphical representations [41]. The same is worth noting for the various systemic diseases herein presented, as the majority have suffered changes on their diagnosis or disease staging, contributing to high heterogeneity and possible unstable meta-analytical statistical power and significance.

In what meta-analyses from intervention studies concern, the short follow-up studies in overall oral research is also a major restraint, previously discussed [13,42]. As a result of the knowledge obtained from the impact of periodontal therapies on systemic inflammation [33] and glycemia [43], these outcomes may occur several months postintervention, thus oral research community shall expand the follow-up longer to allow a more convincing and lasting outlook.

### Implications for Practice and Research

The relatively high number of meta-analyses included roots the notion that the links between oral conditions and systemic NCDs have been considerably increasing. While the overall evidence consistency is unfavorable due to poor meta-analytic or methodological reasons, these results substantiated significant associations with systemic NCDs that are importantly prevalent, such as diabetes mellitus, CVDs, rheumatic or neurodegenerative conditions. This does not mean that other robust associations cannot endure, yet the current body of evidence does not support such inferences. Until the level of evidence becomes clearer, oral care providers shall focus on these particular patients through preventive programs for early detection, oral health literacy promotion, using reliable diagnostic approaches and implementing adequate treatments. Furthermore, all interesting parties shall contribute to a global view of health and not dividing the mouth from the body, and this will foster a growing multidisciplinary care of the patient within its specifics and particularities. Enriching the curricula of medical and dental health courses is an initiative with conceivable impact in the long-term, along with dedicated training towards this unique one health vision, which will naturally expand the observant radar for the impact of oral status to systemic health and vice-versa. These results also enhance the available information with evidence strength endorsements necessary for engaging policy-makers towards increasingly refined programs that will be adapted to the most current evidence.

## Methods

### Protocol and reporting

The protocol was defined a priori by all authors, published online in the PROSPERO platform (ID: CRD42022300740) and performed following the PRISMA guideline [44] (Supplementary Data 6).

### Study selection

For this umbrella review, five electronic databases (PubMed, Cochrane Database of Systematic Reviews, EMBASE, Web of Science, and LILACS) were searched up to December 2021. We merged keywords and subject headings appropriately for each database using the following syntax: (periodontal disease[MeSH] OR oral health[MeSH] OR dental Caries[MeSH] OR “oral manifestations”) AND “systematic review”. In addition, grey literature was searched via http://www.opengrey.eu. Additional relevant literature was included after a manual search of the reference lists of the final included articles. The electronic database search was carried out by two independent authors (J.B. and V.M.), and the final decision for inclusion was made according to the following criteria: (1) systematic reviews with meta-analysis; (2) results from human studies; (3) assessing the association between oral and systemic conditions. There were no restrictions regarding year nor language of publication. As such, exclusion criteria were as follows: (1) systematic reviews without meta-analysis were excluded as it prevented the quantification of the meta-analytical quality of the estimates; (2) systematic reviews reporting binary results without controls; (3) systematic reviews failing to provide meta-analytic estimates and heterogeneity results; (4) systematic reviews of systematic reviews (umbrella reviews).

### Data extraction

We prepared a predefined table to extract the necessary data from each eligible systematic review, including: study identification (authors and year), number of studies included in the meta-analysis, type and number of studies included, oral condition(s) being assessed, systemic condition(s) being assessed, methodological quality tool used, effect size and 95% CI, funding information. From each eligible systematic review, three independent researchers (J.B., V.M., J.V.) extracted information and all disagreements were resolved through discussion with a fourth reviewer (J.J.M.). The agreement between the examiners was considered excellent (0.88, 95% CI: 0.86-0.90).

### Methodological quality appraisal

The included systematic reviews were independently assessed by two examiners (J.B. and V.M.) using the AMSTAR 2 [36]. In this sense, systematic reviews are categorized as: High (“Zero or one non-critical weakness”); Moderate (“More than one non-critical weakness”); Low (“One critical flaw with or without non-critical weaknesses”); and Critically Low (“More than one critical flaw with or without non-critical weaknesses”).

### Grading of the evidence

We graded meta-analyses following a previously published methodology [34]. Significant associations were categorized into four evidence levels: strong, highly suggestive, suggestive, and weak evidence [34,45]. A category of strong evidence was attributed if all the following criteria were met: >1000 cases included in the meta-analysis, a threshold that provides 80% power for hazard ratios ≥1.20 (α=0.05) [34]; a P value ≤10^−6^ of statistical significance in valid meta-analysis [46–48]; heterogeneity (I^2^) below 50%; the null value was excluded by the 95% prediction interval; and, no evidence of small study effects and excess significance bias. Highly suggestive evidence was set if: meta-analyses with >1000 cases; a random effects P value ≤10^−6^, and the largest study in the meta-analysis was statistically significant. Suggestive evidence was defined if: meta-analyses with >1000 cases, random-effects ≤10^−3^ [46–48] were categorized. If the latter conditions were not verified, the meta-analysis was categorized as of weak evidence.

### Calculation of FSN (fail-safe number)

In nominally statistically significant meta-analyses, we determined the number of future studies of average null effect and average weight needed to detect a non-statistically significant summary estimate by calculating Rosenberg’s FSN [49]. Using the Meta-Essentials packages for binary (odds ratio, risk ratio, hazard ratio, incidence ratio or ratio of means) and continuous measures (mean difference, standardized mean difference or weighted mean difference) [50]. We then calculated the median and range for each evidence grade (strong, highly suggestive, suggestive and weak).

### Data handling and management

All data were collected in MS Office 365. Inferential statistical analyses were computed using R version 4.03.

## Data Availability

All data produced in the present study are available upon reasonable request to the authors

## Acknowledgements

This work is financed by national funds through the FCT—Foundation for Science and Technology, I.P., under the Project UIDB/04585/2020. The study sponsor had no role in the design and conduct of the study; collection, management, analysis and interpretation of the data; preparation, review or approval of the article; and decision to submit the article for publication.

## Author contributions

The study was conceived and designed by J.B. The data were acquired and collated by J.B., V.M., and J.V. and analyzed by J.B., and V.M. The manuscript was drafted by J.B. and revised critically for important intellectual content by J.B., P.M., J.V., L.P., M.O., Y.L., L.C., J.J.M., and V.M. All authors gave final approval of the version to be published and have contributed to the manuscript.

## Competing interests

The authors declare no competing interests.

## Notes

### Competing Interest Statement

The authors have declared no competing interest.

