## Supplementary material for "An umbrella review of the evidence linking oral health and systemic health: from the prevalence to clinical and circulating markers": Supplementary data 1.docx

**Supplementary Data 1.** List of excluded studies with justification for exclusion.

| n.º | Reference | Reason |
| --- | --- | --- |
| 1 | Torlińska-Walkowiak N, Majewska KA, Kędzia A, Opydo-Szymaczek J. J Clin Med. 2021 Aug 22;10(16):3733. doi: 10.3390/jcm10163733. | No Meta-Analysis |
| 2 | Almoosawy SA, McGowan M, Hijazi K, Patey R, Bachoo P, Cherukara G. Vascular. 2021 Aug;29(4):556-566. doi: 10.1177/1708538120963914. Epub 2020 Oct 12. | No control group |
| 3 | Sadeghi M, Golshah A, Godiny M, Sharifi R, Khavid A, Nikkerdar N, Tadakamadla SK. Children (Basel). 2021 Apr 15;8(4):302. doi: 10.3390/children8040302. | Unrelated |
| 4 | Agossa K, Roman L, Gosset M, Yzet C, Fumery M. Expert Rev Gastroenterol Hepatol. 2021 Jul 19:1-15. doi: 10.1080/17474124.2021.1952866. Online ahead of print. | No control group |
| 5 | Mahdi SS, Jafri HA, Allana R, Amenta F, Khawaja M, Qasim SSB. Int J Environ Res Public Health. 2021 Jan 28;18(3):1162. doi: 10.3390/ijerph18031162. | No control group |
| 6 | Difloe-Geisert JC, Bernauer SA, Schneeberger N, Bornstein MM, Walter C. Clin Oral Investig. 2021 Jun;25(6):3341-3349. doi: 10.1007/s00784-021-03873-0. Epub 2021 Mar 22. | No control group |
| 7 | Shuai Y, Liu B, Zhou G, Rong L, Niu C, Jin L. Oral Dis. 2021 Oct;27(7):1616-1620. doi: 10.1111/odi.13549. Epub 2020 Aug 2. | Unrelated |
| 8 | Meurman JH, Bascones-Martinez A. Oral Health Prev Dent. 2021 Jan 7;19(1):441-448. doi: 10.3290/j.ohpd.b1993965. | No Meta-Analysis |
| 9 | Apessos I, Voulgaris A, Agrafiotis M, Andreadis D, Steiropoulos P. BMC Pulm Med. 2021 Mar 18;21(1):92. doi: 10.1186/s12890-021-01429-2. | No Meta-Analysis |
| 10 | Khadka S, Khan S, King A, Goldberg LR, Crocombe L, Bettiol S. Age Ageing. 2021 Jan 8;50(1):81-87. doi: 10.1093/ageing/afaa102. | No Meta-Analysis |
| 11 | Jang H, Patoine A, Wu TT, Castillo DA, Xiao J. Sci Rep. 2021 Aug 19;11(1):16870. doi: 10.1038/s41598-021-96495-1. | Unrelated |
| 12 | Thomson WM, Barak Y. J Dent Res. 2021 Mar;100(3):226-231. doi: 10.1177/0022034520957233. Epub 2020 Sep 18. | Not a Systematic Review |
| 13 | Kelly N, Winning L, Irwin C, Lundy FT, Linden D, McGarvey L, Linden GJ, El Karim IA. BMC Oral Health. 2021 Sep 3;21(1):425. doi: 10.1186/s12903-021-01757-z. | No Meta-Analysis |
| 14 | Rajadurai SG, Maharajan MK, Veettil SK, Gopinath D. J Fungi (Basel). 2021 Aug 5;7(8):637. doi: 10.3390/jof7080637. | Unrelated |
| 15 | Taylor HL, Rahurkar S, Treat TJ, Thyvalikakath TP, Schleyer TK. J Dent Res. 2021 Mar;100(3):253-260. doi: 10.1177/0022034520965958. Epub 2020 Oct 22. | No Meta-Analysis |
| 16 | Alvarenga MOP, Frazão DR, de Matos IG, Bittencourt LO, Fagundes NCF, Rösing CK, Maia LC, Lima RR. Front Aging Neurosci. 2021 May 24;13:651437. doi: 10.3389/fnagi.2021.651437. eCollection 2021. | No Meta-Analysis |
| 17 | Sadaf D, Ahmad MZ. J Evid Based Dent Pract. 2021 Sep;21(3):101616. doi: 10.1016/j.jebdp.2021.101616. Epub 2021 Jul 16. | Commentary |
| 18 | Alvarenga MOP, Miranda GHN, Ferreira RO, Saito MT, Fagundes NCF, Maia LC, Lima RR. Front Public Health. 2021 Jan 8;8:550614. doi: 10.3389/fpubh.2020.550614. eCollection 2020. | No Meta-Analysis |
| 19 | Hujoel PP. J Evid Based Dent Pract. 2021 Jun;21(2):101534. doi: 10.1016/j.jebdp.2021.101534. Epub 2021 Feb 23. | Commentary |
| 20 | de Mello-Neto JM, Nunes JGR, Tadakamadla SK, da Silva Figueredo CM. Int J Environ Res Public Health. 2021 Aug 25;18(17):8958. doi: 10.3390/ijerph18178958. | No Meta-Analysis |
| 21 | Lanau N, Mareque-Bueno J, Zabalza M. Eur J Dent. 2021 Feb;15(1):168-173. doi: 10.1055/s-0040-1718244. Epub 2020 Oct 8. | No Meta-Analysis |
| 22 | Lecaplain B, Badran Z, Soueidan A, Prud'homme T, Gaudin A. Andrology. 2021 May;9(3):769-780. doi: 10.1111/andr.12961. Epub 2021 Jan 2. | No Meta-Analysis |
| 23 | Lafuente-Ibáñez de Mendoza I, Cayero-Garay A, Quindós-Andrés G, Aguirre-Urizar JM. Int J Implant Dent. 2021 Jun 17;7(1):73. doi: 10.1186/s40729-021-00338-7. | Unrelated |
| 24 | Costa MJF, de Araújo IDT, da Rocha Alves L, da Silva RL, Dos Santos Calderon P, Borges BCD, de Aquino Martins ARL, de Vasconcelos Gurgel BC, Lins RDAU. Clin Oral Investig. 2021 Mar;25(3):797-806. doi: 10.1007/s00784-020-03764-w. Epub 2021 Jan 20. | No Meta-Analysis |
| 25 | Mizutani K, Buranasin P, Mikami R, Takeda K, Kido D, Watanabe K, Takemura S, Nakagawa K, Kominato H, Saito N, Hattori A, Iwata T. Antioxidants (Basel). 2021 Aug 18;10(8):1304. doi: 10.3390/antiox10081304. | Unrelated |
| 26 | Jakovljevic A, Sljivancanin Jakovljevic T, Duncan HF, Nagendrababu V, Jacimovic J, Aminoshariae A, Milasin J, Dummer PMH. Int Endod J. 2021 Sep;54(9):1527-1537. doi: 10.1111/iej.13538. Epub 2021 May 28. | No Meta-Analysis |
| 27 | Charlotte Höfer K, Graf I, Adams A, Kuhr K, Plum G, Schwendicke F, Brockmeier K, Johannes Noack M. Oral Dis. 2021 Jul 10. doi: 10.1111/odi.13957. Online ahead of print. | No control group |
| 28 | Fageeh HN, Fageeh HI, Prabhu A, Bhandi S, Khan S, Patil S. Syst Rev. 2021 Jan 4;10(1):5. doi: 10.1186/s13643-020-01554-9. | Unrelated |
| 29 | Al-Maweri SA, Ibraheem WI, Al-Ak'hali MS, Shamala A, Halboub E, Alhajj MN. Crit Rev Oncol Hematol. 2021 Mar;159:103221. doi: 10.1016/j.critrevonc.2021.103221. Epub 2021 Jan 20. | No Meta-Analysis |
| 30 | Casillas Santana MA, Arreguín Cano JA, Dib Kanán A, Dipp Velázquez FA, Munguía PDCS, Martínez Castañón GA, Castillo Silva BE, Sámano Valencia C, Salas Orozco MF. Medicina (Kaunas). 2021 May 13;57(5):493. doi: 10.3390/medicina57050493. | No Meta-Analysis |
| 31 | Muniz FWMG, Pola NM, Silva CFE, Silva FGD, Casarin M. Arch Oral Biol. 2021 Sep;129:105184. doi: 10.1016/j.archoralbio.2021.105184. Epub 2021 Jun 6. | No Meta-Analysis |
| 32 | Contaldo M, Lucchese A, Romano A, Della Vella F, Di Stasio D, Serpico R, Petruzzi M. Int J Mol Sci. 2021 Aug 26;22(17):9251. doi: 10.3390/ijms22179251. | No Meta-Analysis |
| 33 | Borsa L, Dubois M, Sacco G, Lupi L. Int J Environ Res Public Health. 2021 Sep 3;18(17):9312. doi: 10.3390/ijerph18179312. | No Meta-Analysis |
| 34 | Delbove T, Gueyffier F, Juillard L, Kalbacher E, Maucort-Boulch D, Nony P, Grosgogeat B, Gritsch K. PLoS One. 2021 Jan 22;16(1):e0245619. doi: 10.1371/journal.pone.0245619. eCollection 2021. | No Meta-Analysis |
| 35 | Lembo D, Caroccia F, Lopes C, Moscagiuri F, Sinjari B, D'Attilio M. Medicina (Kaunas). 2021 Jun 21;57(6):640. doi: 10.3390/medicina57060640. | No Meta-Analysis |
| 36 | Plemmenos G, Evangeliou E, Polizogopoulos N, Chalazias A, Deligianni M, Piperi C. Curr Med Chem. 2021;28(15):3032-3058. doi: 10.2174/0929867327666200824112732. | Unrelated |
| 37 | Shelswell J. Evid Based Dent. 2021 Jan;22(1):40-41. doi: 10.1038/s41432-021-0156-4. | Commentary |
| 38 | Bhola S, Geddis-Regan A. Evid Based Dent. 2021 Jan;22(1):14-15. doi: 10.1038/s41432-021-0149-3. | Commentary |
| 39 | Corbella S, Calciolari E, Alberti A, Donos N, Francetti L. Sci Rep. 2021 Jun 9;11(1):12125. doi: 10.1038/s41598-021-91506-7. | Unrelated |
| 40 | Cuevas-Gonzalez MV, Espinosa-Cristóbal LF, Donohue-Cornejo A, Tovar-Carrillo KL, Saucedo-Acuña RA, García-Calderón AG, Guzmán-Gastelum DA, Cuevas-Gonzalez JC. Medicine (Baltimore). 2021 Dec 23;100(51):e28327. doi: 10.1097/MD.0000000000028327. | No Meta-Analysis |
| 41 | Zupo R, Castellana F, De Nucci S, Dibello V, Lozupone M, Giannelli G, De Pergola G, Panza F, Sardone R, Boeing H. Front Nutr. 2021 Oct 27;8:762383. doi: 10.3389/fnut.2021.762383. eCollection 2021. | No Meta-Analysis |
| 42 | Algra Y, Haverkort E, Kok W, Etten-Jamaludin FV, Schoot LV, Hollaar V, Naumann E, Schueren MV, Jerković-Ćosić K. Nutrients. 2021 Oct 13;13(10):3584. doi: 10.3390/nu13103584. | No Meta-Analysis |
| 43 | Nascimento RB, Araujo NS, Silva JC, Xavier FCA. Spec Care Dentist. 2021 Nov 18:10.1111/scd.12669. doi: 10.1111/scd.12669. Online ahead of print. | No Meta-Analysis |
| 44 | Nijakowski K, Gruszczyński D, Surdacka A. Int J Environ Res Public Health. 2021 Nov 2;18(21):11521. doi: 10.3390/ijerph182111521. | Failed to provide heterogeneity values |
| 45 | Macnamara A, Mishu MP, Faisal MR, Islam M, Peckham E. PLoS One. 2021 Dec 1;16(12):e0260766. doi: 10.1371/journal.pone.0260766. eCollection 2021. | No Meta-Analysis |
| 46 | Elwishahy A, Antia K, Bhusari S, Ilechukwu NC, Horstick O, Winkler V. J Alzheimers Dis Rep. 2021 Sep 13;5(1):721-732. doi: 10.3233/ADR-200237. eCollection 2021. | No Meta-Analysis |
| 47 | Reitano E, de'Angelis N, Gavriilidis P, Gaiani F, Memeo R, Inchingolo R, Bianchi G, de'Angelis GL, Carra MC. Microorganisms. 2021 Dec 14;9(12):2585. doi: 10.3390/microorganisms9122585. | No Meta-Analysis |
| 48 | Guerrero-Gironés J, Ros-Valverde A, Pecci-Lloret MP, Rodríguez-Lozano FJ, Pecci-Lloret MR. J Clin Med. 2021 Oct 23;10(21):4886. doi: 10.3390/jcm10214886. | No Meta-Analysis |
| 49 | Parra-Torres V, Melgar-Rodríguez S, Muñoz-Manríquez C, Sanhueza B, Cafferata EA, Paula-Lima AC, Díaz-Zúñiga J. Oral Dis. 2021 Oct 26. doi: 10.1111/odi.14054. Online ahead of print. | No Meta-Analysis |
| 50 | Contaldo M, Santoro R, Romano A, Loffredo F, Di Stasio D, Della Vella F, Scivetti M, Petruzzi M, Serpico R, Lucchese A. Oral Manifestations in Children and Young Adults with Down Syndrome: A Systematic Review of the Literature. Applied Sciences. 2021; 11(12):5408. <https://doi.org/10.3390/app11125408> | No Meta-Analysis |
| 51 | Elwishahy A, Antia K, Bhusari S, Ilechukwu NC, Horstick O, Winkler V. Porphyromonas Gingivalis as a Risk Factor to Alzheimer's Disease: A Systematic Review. J Alzheimers Dis Rep. 2021 Sep 13;5(1):721-732. doi: 10.3233/ADR-200237. PMID: 34755046; PMCID: PMC8543378. | No Meta-Analysis |
| 52 | Raju K, Taylor GW, Tahir P, Hyde S. Association of tooth loss with morbidity and mortality by diabetes status in older adults: a systematic review. BMC Endocr Disord. 2021 Oct 19;21(1):205. doi: 10.1186/s12902-021-00830-6. PMID: 34663281; PMCID: PMC8524900. | No Meta-Analysis |
| 53 | Mester A, Mancini L, Marchetti E, Baciut M, Bran S, Lucaciu O, Baciut G, Tomuleasa C, Pasca S, Piciu A, Voina-Tonea A, Opris H, Prodan DA, Onisor F. The Presence of Periodontitis in Patients with Von Willebrand Disease: A Systematic Review. Applied Sciences. 2021; 11(14):6408. <https://doi.org/10.3390/app11146408> | No Meta-Analysis |
| 54 | Chu XJ, Cao NW, Zhou HY, Meng X, Guo B, Zhang HY, Li BZ. The oral and gut microbiome in rheumatoid arthritis patients: a systematic review. Rheumatology (Oxford). 2021 Mar 2;60(3):1054-1066. doi: 10.1093/rheumatology/keaa835. PMID: 33450018. | No Meta-Analysis |
| 55 | Santoso CMA, Ketti F, Bramantoro T, Zsuga J, Nagy A. Association between Oral Hygiene and Metabolic Syndrome: A Systematic Review and Meta-Analysis. J Clin Med. 2021 Jun 28;10(13):2873. doi: 10.3390/jcm10132873. PMID: 34203460; PMCID: PMC8269064. | No clinical measures |
| 56 | Corridore D, Zumbo G, Corvino I, Guaragna M, Bossù M, Polimeni A, Vozza I. Clin Ter. 2020 May-Jun;171(3):e275-e282. doi: 10.7417/CT.2020.2226. | No Meta-Analysis |
| 57 | Carvalho Silva C, Mendes R, Manso MDC, Gavinha S, Melo P. Oral Health Prev Dent. 2020 Sep 4;18(1):653-667. doi: 10.3290/j.ohpd.a45089. | Unrelated |
| 58 | Manchery N, Henry JD, Nangle MR. Community Dent Oral Epidemiol. 2020 Apr;48(2):89-100. doi: 10.1111/cdoe.12512. Epub 2019 Dec 9. | No Meta-Analysis |
| 59 | Angst PDM, Maier J, Dos Santos Nogueira R, Manso IS, Tedesco TK. Arch Oral Biol. 2020 Dec;120:104948. doi: 10.1016/j.archoralbio.2020.104948. Epub 2020 Oct 16. | No control group |
| 60 | Vandeplas C, Vranckx M, Hekner D, Politis C, Jacobs R. J Oral Maxillofac Surg. 2020 Nov;78(11):1892-1908. doi: 10.1016/j.joms.2020.06.014. Epub 2020 Jun 15. | No Meta-Analysis |
| 61 | Lechien JR, Chiesa-Estomba CM, Calvo Henriquez C, Mouawad F, Ristagno C, Barillari MR, Schindler A, Nacci A, Bouland C, Laino L, Saussez S. PLoS One. 2020 Aug 14;15(8):e0237581. doi: 10.1371/journal.pone.0237581. eCollection 2020. | No Meta-Analysis |
| 62 | Cagetti MG, Wolf TG, Tennert C, Camoni N, Lingström P, Campus G. Int J Environ Res Public Health. 2020 Feb 3;17(3):938. doi: 10.3390/ijerph17030938. | Unrelated |
| 63 | Castrillón E, Castro C, Ojeda A, Caicedo N, Moreno S, Moreno F. Rev Colomb Psiquiatr (Engl Ed). 2020 Jun 2:S0034-7450(20)30026-3. doi: 10.1016/j.rcp.2020.02.001. Online ahead of print. | No control group |
| 64 | Moosazadeh M, Gorji NE, Nasiri P, Shafaroudi AM. BDJ Open. 2020 Nov 17;6(1):23. doi: 10.1038/s41405-020-00051-4. | Restricted to a particular country |
| 65 | Indrastiti RK, Wardhany II, Soegyanto AI. Oral Dis. 2020 Sep;26 Suppl 1:133-136. doi: 10.1111/odi.13394. | No Meta-Analysis |
| 66 | Coffey N, O' Leary F, Burke F, Roberts A, Hayes M. J Dent. 2020 Dec;103:103509. doi: 10.1016/j.jdent.2020.103509. Epub 2020 Oct 28. | Abstract |
| 67 | Lechien JR, Calvo-Henriquez C, Chiesa-Estomba CM, Barillari MR, Trozzi M, Meucci D, Peer S, Ben Abdelouahed F, Schindler A, Saussez S. Int J Pediatr Otorhinolaryngol. 2020 Sep;136:110166. doi: 10.1016/j.ijporl.2020.110166. Epub 2020 Jun 8. | No Meta-Analysis |
| 68 | Siddiqi A, Zafar S, Sharma A, Quaranta A. J Interprof Care. 2020 Dec 8:1-9. doi: 10.1080/13561820.2020.1825354. Online ahead of print. | No Meta-Analysis |
| 69 | Nguyen ATM, Akhter R, Garde S, Scott C, Twigg SM, Colagiuri S, Ajwani S, Eberhard J. Diabetes Res Clin Pract. 2020 Jul;165:108244. doi: 10.1016/j.diabres.2020.108244. Epub 2020 Jun 8. | No Meta-Analysis |
| 70 | Lam PP, Du R, Peng S, McGrath CP, Yiu CK. Autism. 2020 Jul;24(5):1047-1066. doi: 10.1177/1362361319877337. Epub 2020 Jan 13. | No Meta-Analysis |
| 71 | Mester A, Irimie AI, Tanase A, Tranca S, Campian RS, Tomuleasa C, Dima D, Piciu A, Lucaciu O. Crit Rev Oncol Hematol. 2020 Mar;147:102878. doi: 10.1016/j.critrevonc.2020.102878. Epub 2020 Jan 23. | No Meta-Analysis |
| 72 | Venkatasalu MR, Murang ZR, Ramasamy DTR, Dhaliwal JS. BMC Oral Health. 2020 Mar 18;20(1):79. doi: 10.1186/s12903-020-01075-w. | No Meta-Analysis |
| 73 | Pandey A, Rajak R, Pandey M. Eur J Rheumatol. 2020 Dec 1. doi: 10.5152/eurjrheum.2020.20177. Online ahead of print. | No Meta-Analysis |
| 74 | Govindasamy R, Periyasamy S, Narayanan M, Balaji VR, Dhanasekaran M, Karthikeyan B. J Indian Soc Periodontol. 2020 Jan-Feb;24(1):7-14. doi: 10.4103/jisp.jisp_228_19. Epub 2020 Jan 2. | No Meta-Analysis |
| 75 | Pérez-Losada FL, Estrugo-Devesa A, Castellanos-Cosano L, Segura-Egea JJ, López-López J, Velasco-Ortega E. J Clin Med. 2020 Feb 17;9(2):540. doi: 10.3390/jcm9020540. | No Meta-Analysis |
| 76 | Orlandini RK, Bepu DAN, Saraiva MDCP, Bollela VR, Motta ACF, Lourenço AG. Microb Pathog. 2020 Dec;149:104477. doi: 10.1016/j.micpath.2020.104477. Epub 2020 Sep 10. | Unrelated |
| 77 | Nagendrababu V, Segura-Egea JJ, Fouad AF, Pulikkotil SJ, Dummer PMH. Int Endod J. 2020 Apr;53(4):455-466. doi: 10.1111/iej.13253. Epub 2019 Dec 19. | No Meta-Analysis |
| 78 | Borra V, Darius A, Dockx K, Compernolle V, Lambrechts P, Vandekerckhove P, De Buck E. Int J Evid Based Healthc. 2020 Jun;18(2):170-187. doi: 10.1097/XEB.0000000000000219. | Unrelated |
| 79 | Jakovljevic A, Duncan HF. J Evid Based Dent Pract. 2020 Sep;20(3):101467. doi: 10.1016/j.jebdp.2020.101467. Epub 2020 Jun 17. | Commentary |
| 80 | da Costa Lopes AJ, Cunha TCA, Monteiro MCM, Serra-Negra JM, Cabral LC, Júnior PCS. Sleep Breath. 2020 Sep;24(3):913-921. doi: 10.1007/s11325-019-01919-y. Epub 2019 Oct 18. | No Meta-Analysis |
| 81 | Pulikkotil SJ, Nath S; Muthukumaraswamy, Dharamarajan L, Jing KT, Vaithilingam RD. Community Dent Health. 2020 Feb 27;37(1):12-21. doi: 10.1922/CDH_4569Pulikkotil10. | Unrelated |
| 82 | Schmidlin PR, Khademi A, Fakheran O. Clin Oral Investig. 2020 Oct;24(10):3335-3345. doi: 10.1007/s00784-020-03475-2. Epub 2020 Jul 31. | No Meta-Analysis |
| 83 | Nunes-Dos-Santos DL, Gomes SV, Rodrigues VP, Pereira ALA. Oral Dis. 2020 Jan;26(1):22-34. doi: 10.1111/odi.13040. Epub 2019 Feb 6. | No Meta-Analysis |
| 84 | Ayilavarapu S. J Evid Based Dent Pract. 2020 Jun;20(2):101412. doi: 10.1016/j.jebdp.2020.101412. Epub 2020 Feb 20. | Commentary |
| 85 | Moosavi MS, Barati H. Acta Clin Belg. 2020 Feb;75(1):19-25. doi: 10.1080/17843286.2018.1540164. Epub 2018 Oct 30. | Unsuitable inclusion criteria |
| 86 | Suresh Unniachan A, Krishnavilasom Jayakumari N, Sethuraman S. Curr Med Mycol. 2020 Jun;6(2):63-68. doi: 10.18502/CMM.6.2.3420. | Unrelated |
| 87 | Kelly N, El Karim I. J Evid Based Dent Pract. 2020 Dec;20(4):101498. doi: 10.1016/j.jebdp.2020.101498. Epub 2020 Sep 29. | No Meta-Analysis |
| 88 | Kim JY, Kim HN. Int J Environ Res Public Health. 2020 Dec 29;18(1):194. doi: 10.3390/ijerph18010194. | Unrelated |
| 89 | Konopka T, Zakrzewska A. Ginekol Pol. 2020;91(3):158-164. doi: 10.5603/GP.2020.0024. | No Meta-Analysis |
| 90 | Jakovljevic A, Duncan HF, Nagendrababu V, Jacimovic J, Milasin J, Dummer PMH. Int Endod J. 2020 Oct;53(10):1374-1386. doi: 10.1111/iej.13364. Epub 2020 Aug 8. | No Meta-Analysis |
| 91 | Dreweck FDS, Soares S, Duarte J, Conti PCR, De Luca Canto G, Luís Porporatti A. J Oral Rehabil. 2020 Aug;47(8):1041-1051. doi: 10.1111/joor.12993. Epub 2020 May 26. | No Meta-Analysis |
| 92 | Froum SJ, Hengjeerajaras P, Liu KY, Maketone P, Patel V, Shi Y. Int J Periodontics Restorative Dent. 2020 Nov/Dec;40(6):e229-e233. doi: 10.11607/prd.4591. | No Meta-Analysis |
| 93 | Samborska-Mazur J, Sikorska D, Wyganowska-Świątkowska M. Reumatologia. 2020;58(4):236-242. doi: 10.5114/reum.2020.98436. Epub 2020 Aug 31. | No Meta-Analysis |
| 94 | Zamri F, de Vries TJ. Front Immunol. 2020 Oct 23;11:591365. doi: 10.3389/fimmu.2020.591365. eCollection 2020. | No Meta-Analysis |
| 95 | Machado V, Lobo S, Proença L, Mendes JJ, Botelho J. Nutrients. 2020 Jul 22;12(8):2177. doi: 10.3390/nu12082177. | Unrelated |
| 96 | Lafuente Ibáñez de Mendoza I, Maritxalar Mendia X, García de la Fuente AM, Quindós Andrés G, Aguirre Urizar JM. J Periodontal Res. 2020 Jan;55(1):13-22. doi: 10.1111/jre.12691. Epub 2019 Sep 17. | No Meta-Analysis |
| 97 | Colonia-García A, Gutiérrez-Vélez M, Duque-Duque A, de Andrade CR. Acta Odontol Scand. 2020 Oct;78(7):553-559. doi: 10.1080/00016357.2020.1774076. Epub 2020 Jun 18. | No Meta-Analysis |
| 98 | Saravi B, Lang G, Ülkümen S, Burchard T, Weihrauch V, Patzelt S, Boeker M, Li Z, Woelber JP. Eur Cell Mater. 2020 Nov 9;40:203-226. doi: 10.22203/eCM.v040a13. | No Meta-Analysis |
| 99 | Brum RS, Duarte PM, Canto GL, Flores-Mir C, Benfatti CAM, Porporatti AL, Zimmermann GS. J Indian Soc Periodontol. 2020 May-Jun;24(3):191-215. doi: 10.4103/jisp.jisp_512_19. Epub 2020 May 4. | Failed to provide heterogeneity values |
| 100 | Kruse AB, Kowalski CD, Leuthold S, Vach K, Ratka-Krüger P, Woelber JP. Lipids Health Dis. 2020 May 21;19(1):100. doi: 10.1186/s12944-020-01267-x. | Unrelated |
| 101 | Koidou VP, Chatzopoulos GS, Tomas I, Nibali L, Donos N. Clin Oral Investig. 2020 Jan;24(1):487-502. doi: 10.1007/s00784-019-03088-4. Epub 2019 Nov 7. | No Meta-Analysis |
| 102 | Né YGS, Martins BV, Castro MML, Alvarenga MOP, Fagundes NCF, Magno MB, Maia LC, Lima RR. Clin Nutr. 2020 Sep;39(9):2639-2646. doi: 10.1016/j.clnu.2019.12.016. Epub 2019 Dec 28. | No Meta-Analysis |
| 103 | Cotti E, Cairo F, Bassareo PP, Fonzar F, Venturi M, Landi L, Parolari A, Franco V, Fabiani C, Barili F, Di Lenarda A, Gulizia M, Borzi M, Campus G, Musumeci F, Mercuro G. Int Endod J. 2020 Feb;53(2):186-199. doi: 10.1111/iej.13166. Epub 2019 Dec 20. | No Meta-Analysis |
| 104 | Koidou VP, Cavalli N, Hagi-Pavli E, Nibali L, Donos N. J Periodontal Res. 2020 Dec;55(6):801-809. doi: 10.1111/jre.12795. Epub 2020 Aug 25. | No Meta-Analysis |
| 105 | O'Connor JP, Milledge KL, O'Leary F, Cumming R, Eberhard J, Hirani V. Nutr Rev. 2020 Feb 1;78(2):175-188. doi: 10.1093/nutrit/nuz035. | No Meta-Analysis |
| 106 | Decker A, Askar H, Tattan M, Taichman R, Wang HL. Clin Oral Investig. 2020 Jan;24(1):1-12. doi: 10.1007/s00784-019-03089-3. Epub 2019 Nov 1. | No Meta-Analysis |
| 107 | Márquez-Arrico CF, Silvestre-Rangil J, Gutiérrez-Castillo L, Martinez-Herrera M, Silvestre FJ, Rocha M. J Clin Med. 2020 May 23;9(5):1586. doi: 10.3390/jcm9051586. | No Meta-Analysis |
| 108 | Nardi GM, Ferrara E, Converti I, Cesarano F, Scacco S, Grassi R, Gnoni A, Grassi FR, Rapone B. Int J Environ Res Public Health. 2020 Apr 16;17(8):2765. doi: 10.3390/ijerph17082765. | No Meta-Analysis |
| 109 | Bogdan M, Meca AD, Boldeanu MV, Gheorghe DN, Turcu-Stiolica A, Subtirelu MS, Boldeanu L, Blaj M, Botnariu GE, Vlad CE, Foia LG, Surlin P. Nutrients. 2020 Feb 20;12(2):553. doi: 10.3390/nu12020553. | Unrelated |
| 110 | Dioguardi M, Crincoli V, Laino L, Alovisi M, Sovereto D, Mastrangelo F, Russo LL, Muzio LL. J Clin Med. 2020 Feb 11;9(2):495. doi: 10.3390/jcm9020495. | No Meta-Analysis |
| 111 | Chinnasamy A, Moodie M. Int J Dent. 2020 Aug 25;2020:2964020. doi: 10.1155/2020/2964020. eCollection 2020. | Unrelated |
| 112 | Willeit P, Tschiderer L, Allara E, Reuber K, Seekircher L, Gao L, Liao X, Lonn E, Gerstein HC, Yusuf S, Brouwers FP, Asselbergs FW, van Gilst W, Anderssen SA, Grobbee DE, Kastelein JJP, Visseren FLJ, Ntaios G, Hatzitolios AI, Savopoulos C, Nieuwkerk PT, Stroes E, Walters M, Higgins P, Dawson J, Gresele P, Guglielmini G, Migliacci R, Ezhov M, Safarova M, Balakhonova T, Sato E, Amaha M, Nakamura T, Kapellas K, Jamieson LM, Skilton M, Blumenthal JA, Hinderliter A, Sherwood A, Smith PJ, van Agtmael MA, Reiss P, van Vonderen MGA, Kiechl S, Klingenschmid G, Sitzer M, Stehouwer CDA, Uthoff H, Zou ZY, Cunha AR, Neves MF, Witham MD, Park HW, Lee MS, Bae JH, Bernal E, Wachtell K, Kjeldsen SE, Olsen MH, Preiss D, Sattar N, Beishuizen E, Huisman MV, Espeland MA, Schmidt C, Agewall S, Ok E, Aşçi G, de Groot E, Grooteman MPC, Blankestijn PJ, Bots ML, Sweeting MJ, Thompson SG, Lorenz MW; PROG-IMT and the Proof-ATHERO Study Groups. Circulation. 2020 Aug 18;142(7):621-642. doi: 10.1161/CIRCULATIONAHA.120.046361. Epub 2020 Jun 17. | Unrelated |
| 113 | Alotaibi WM, et al. Dental Caries in Relation to Obesity in Children: A Systematic Review and Metaanalysis. Ann Med Health Sci Res. 2020;10: 1029-1033. | No control group |
| 114 | Fagundes NCF, Couto RSD, Brandão APT, Lima LAO, Bittencourt LO, Souza-Rodrigues RD, Freire MAM, Maia LC, Lima RR. Association between Tooth Loss and Stroke: A Systematic Review. J Stroke Cerebrovasc Dis. 2020 Aug;29(8):104873. doi: 10.1016/j.jstrokecerebrovasdis.2020.104873. Epub 2020 Jun 8. PMID: 32689647. | No Meta-Analysis |
| 115 | Kapferer-Seebacher I, Schnabl D, Zschocke J, Pope FM. Dental Manifestations of Ehlers-Danlos Syndromes: A Systematic Review. Acta Derm Venereol. 2020 Mar 25;100(7):adv00092. doi: 10.2340/00015555-3428. PMID: 32147746. | No Meta-Analysis |
| 116 | Pandey A, Ravindran V, Pandey M, et alAB0713 PERIODONTAL DISEASES AND ITS ASSOCIATION WITH ANKYLOSING SPONDYLITIS/SPA: A SYSTEMATIC REVIEWAnnals of the Rheumatic Diseases 2020;79:1652. | Abstract |
| 117 | Li P, Shu Y, Gu Y. The potential role of bacteria in pancreatic cancer: a systematic review. Carcinogenesis. 2020 Jun 17;41(4):397-404. doi: 10.1093/carcin/bgaa013. PMID: 32034405. | No Meta-Analysis |
| 118 | Ramon R, Yahya H, Setiawan MR (2020). Association between chronic periodontitis and risk of erectile dysfunction: a systematic review and meta-analysis. UOP-1309. International Journal of Urology. <https://doi.org/10.1111/iju.14397> | Abstract |
| 119 | Santos Silva D, Costa F, Poiares Baptista I, et alTHU0092 USING EVIDENCE-BASED RESEARCH TO DESIGN A RANDOMISED TRIAL ON PERIODONTAL TREATMENT FOR INDIVIDUALS WITH RHEUMATOID ARTHRITIS: A SYSTEMATIC REVIEW PUTTING EXISTING RESEARCH INTO CONTEXTAnnals of the Rheumatic Diseases 2020;79:259. | Abstract |
| 120 | Brignardello-Petersen R. There is still no high-quality evidence that periodontitis is a risk factor for hypertension or that periodontal treatment has beneficial effects on blood pressure. J Am Dent Assoc. 2020 Apr;151(4):e31. doi: 10.1016/j.adaj.2019.11.002. Epub 2020 Feb 4. PMID: 32033741. | Abstract |
| 121 | Wang Y, Xing L, Yu H, Zhao L. BMC Oral Health. 2019 Sep 14;19(1):213. doi: 10.1186/s12903-019-0903-5. | No control group |
| 122 | Pawlaczyk-Kamieńska T, Borysewicz-Lewicka M, Śniatała R, Batura-Gabryel H, Cofta S. J Cyst Fibros. 2019 Nov;18(6):762-771. doi: 10.1016/j.jcf.2018.11.007. Epub 2018 Nov 23. | No Meta-Analysis |
| 123 | Rosten A, Newton T. Ned Tijdschr Tandheelkd. 2019 Mar;126(3):141-150. doi: 10.5177/ntvt.2019.03.19003. | No Meta-Analysis |
| 124 | Ward LM, Cooper SA, Hughes-McCormack L, Macpherson L, Kinnear D. J Intellect Disabil Res. 2019 Nov;63(11):1359-1378. doi: 10.1111/jir.12632. Epub 2019 May 23. | No Meta-Analysis |
| 125 | Lauritano D, Boccalari E, Di Stasio D, Della Vella F, Carinci F, Lucchese A, Petruzzi M. Diagnostics (Basel). 2019 Jul 15;9(3):77. doi: 10.3390/diagnostics9030077. | No control group |
| 126 | Hakeem FF, Bernabé E, Sabbah W. Gerodontology. 2019 Sep;36(3):205-215. doi: 10.1111/ger.12406. Epub 2019 Apr 26. | No control group |
| 127 | Nangle MR, Riches J, Grainger SA, Manchery N, Sachdev PS, Henry JD. Gerontology. 2019;65(6):659-672. doi: 10.1159/000496730. Epub 2019 Mar 22. | No control group |
| 128 | Wallace K, Shafique S, Piamjariyakul U. Nephrol Nurs J. 2019 Jul-Aug;46(4):375-394. | No control group |
| 129 | Fiorillo L, Cervino G, Laino L, D'Amico C, Mauceri R, Tozum TF, Gaeta M, Cicciù M. Dent J (Basel). 2019 Dec 11;7(4):114. doi: 10.3390/dj7040114. | No control group |
| 130 | Nascimento PC, Castro MML, Magno MB, Almeida APCPSC, Fagundes NCF, Maia LC, Lima RR. Front Neurol. 2019 Apr 24;10:323. doi: 10.3389/fneur.2019.00323. eCollection 2019. | No Meta-Analysis |
| 131 | Al-Shamlan SO, Mohammad M, Papandreou D. Open Access Maced J Med Sci. 2019 Jun 28;7(12):2044-2049. doi: 10.3889/oamjms.2019.539. eCollection 2019 Jun 30. | No Meta-Analysis |
| 132 | Salhi L, Rompen E, Sakalihasan N, Laleman I, Teughels W, Michel JB, Lambert F. Angiology. 2019 Jul;70(6):479-491. doi: 10.1177/0003319718821243. Epub 2018 Dec 30. | No Meta-Analysis |
| 133 | Lajolo C, Favia G, Limongelli L, Tempesta A, Zuppa A, Cordaro M, Vanella I, Giuliani M. Acta Otorhinolaryngol Ital. 2019 Apr;39(2):67-74. doi: 10.14639/0392-100X-2281. | No Meta-Analysis |
| 134 | Liu W, Cao Y, Dong L, Zhu Y, Wu Y, Lv Z, Iheozor-Ejiofor Z, Li C. Cochrane Database Syst Rev. 2019 Dec 31;12(12):CD009197. doi: 10.1002/14651858.CD009197.pub4. | No Meta-Analysis |
| 135 | Natto ZS, Hameedaldain A. J Evid Based Dent Pract. 2019 Jun;19(2):131-139. doi: 10.1016/j.jebdp.2018.12.003. Epub 2019 Jan 2. | No Meta-Analysis |
| 136 | Nanayakkara S, Zhou X. J Evid Based Dent Pract. 2019 Jun;19(2):192-194. doi: 10.1016/j.jebdp.2019.05.014. Epub 2019 May 6. | Commentary |
| 137 | Hu L, Zhou M, Young A, Zhao W, Yan Z. BMC Oral Health. 2019 Jul 10;19(1):140. doi: 10.1186/s12903-019-0841-2. | Unrelated |
| 138 | Osorio Parra MM, Elangovan S, Lee CT. Oral Dis. 2019 Jul;25(5):1265-1276. doi: 10.1111/odi.12979. Epub 2018 Oct 9. | No Meta-Analysis |
| 139 | Benrachadi L, Mohamed Saleh Z, Bouziane A. Presse Med. 2019 Jan;48(1 Pt 1):4-18. doi: 10.1016/j.lpm.2018.12.002. Epub 2019 Jan 18. | No Meta-Analysis |
| 140 | Lima RPE, Belém FV, Abreu LG, Cunha FA, Cota LOM, da Costa JE, Costa FO. Int J Periodontics Restorative Dent. 2019 Jan/Feb;39(1):e1-e10. doi: 10.11607/prd.3866. | No Meta-Analysis |
| 141 | Alebel A, Wondemagegn AT, Tesema C, Kibret GD, Wagnew F, Petrucka P, Arora A, Ayele AD, Alemayehu M, Eshetie S. BMC Infect Dis. 2019 Mar 13;19(1):254. doi: 10.1186/s12879-019-3892-8. | Unrelated |
| 142 | Deschamps-Lenhardt S, Martin-Cabezas R, Hannedouche T, Huck O. Oral Dis. 2019 Mar;25(2):385-402. doi: 10.1111/odi.12834. Epub 2018 Apr 19. | No control group |
| 143 | Kaschwich M, Behrendt CA, Heydecke G, Bayer A, Debus ES, Seedorf U, Aarabi G. Int J Mol Sci. 2019 Jun 15;20(12):2936. doi: 10.3390/ijms20122936. | No Meta-Analysis |
| 144 | Voinescu I, Petre A, Burlibasa M, Oancea L. Maedica (Bucur). 2019 Dec;14(4):384-390. doi: 10.26574/maedica.2019.14.4.384. | No Meta-Analysis |
| 145 | Wen S, Beltrán V, Chaparro A, Espinoza F, Riedemann JP. Rev Med Chil. 2019 Jun;147(6):762-775. doi: 10.4067/S0034-98872019000600762. | No Meta-Analysis |
| 146 | Fakheran O, Khodadadi-Bohlouli Z, Khademi A. Gen Dent. 2019 Mar-Apr;67(2):64-67. | Unrelated |
| 147 | Tada A, Miura H. Int J Environ Res Public Health. 2019 Jul 11;16(14):2472. doi: 10.3390/ijerph16142472. | Unrelated |
| 148 | Kriauciunas A, Gleiznys A, Gleiznys D, Janužis G. Cureus. 2019 May 28;11(5):e4775. doi: 10.7759/cureus.4775. | No Meta-Analysis |
| 149 | Glurich I, Acharya A. Curr Diab Rep. 2019 Nov 6;19(11):121. doi: 10.1007/s11892-019-1228-0. | No Meta-Analysis |
| 150 | Segura-Egea JJ, Cabanillas-Balsera D, Jiménez-Sánchez MC, Martín-González J. Int Endod J. 2019 Jun;52(6):790-802. doi: 10.1111/iej.13079. Epub 2019 Feb 14. | No Meta-Analysis |
| 151 | Wang J, Geng X, Sun J, Zhang S, Yu W, Zhang X, Liu H. Rev Cardiovasc Med. 2019 Jun 30;20(2):81-89. doi: 10.31083/j.rcm.2019.02.52. | No Meta-Analysis |
| 152 | Buwembo W, Munabi IG, Kaddumukasa M, Kiryowa H, Nankya E, Johnson WE, Okello E, Sewankambo N. Open J Stomatol. 2019 Oct;9(10):215-226. doi: 10.4236/ojst.2019.910023. Epub 2019 Oct 10. | Unrelated |
| 153 | Tanaka H, Ihana-Sugiyama N, Sugiyama T, Ohsugi M. J Epidemiol. 2019 Jan 5;29(1):1-10. doi: 10.2188/jea.JE20170155. Epub 2018 Jun 23. | Unrelated |
| 154 | Cotti E, Cairo F, Bassareo PP, Fonzar F, Venturi M, Landi L, Parolari A, Franco V, Fabiani C, Barili F, Di Lenarda A, Gulizia M, Borzi M, Campus G, Musumeci F, Mercuro G. Int J Cardiol. 2019 Oct 1;292:78-86. doi: 10.1016/j.ijcard.2019.06.041. Epub 2019 Jun 17. | No Meta-Analysis |
| 155 | Aljudaibi S, Duane B. Evid Based Dent. 2019 Mar;20(1):18-19. doi: 10.1038/s41432-019-0009-6. | Commentary |
| 156 | Meza Maurício J, Miranda TS, Almeida ML, Silva HD, Figueiredo LC, Duarte PM. Braz Oral Res. 2019 Sep 30;33(suppl 1):e070. doi: 10.1590/1807-3107bor-2019.vol33.0070. eCollection 2019. | No Meta-Analysis |
| 157 | Mushtaq S, Aslam MA, Sajjad E. Association between Respiratory Diseases and Oral Health: A Systematic Review Study., Indo Am. J. P. Sci, 2019; 06(05). | No Meta-Analysis |
| 158 | Bett JVS, Batistella EÂ, Melo G, Munhoz EA, Silva CAB, Guerra ENDS, Porporatti AL, De Luca Canto G. Prevalence of oral mucosal disorders during pregnancy: A systematic review and meta-analysis. J Oral Pathol Med. 2019 Apr;48(4):270-277. doi: 10.1111/jop.12831. Epub 2019 Feb 12. PMID: 30673134. | No control group |
| 159 | AlSaleem, FA; Alsadoon, AW. 2019. The Systematic Review of Endo-Perio Lesion and Uncontrolled Diabetes. Indo American Journal of Pharmaceutical Sciences; 6(1): 2617 | No Meta-Analysis |
| 160 | Castilho AVSS, Foratori-Junior GA, Sales-Peres SHC. Bariatric Surgery Impact on Gastroesophageal Reflux and Dental Wear: A Systematic Review. Arq Bras Cir Dig. 2019 Dec 20;32(4):e1466. doi: 10.1590/0102-672020190001e1466. PMID: 31859919; PMCID: PMC6918764. | No Meta-Analysis |
| 161 | Ungprasert P, Wijarnpreecha K, Cheungpasitporn WAB0732 THE ASSOCIATION BETWEEN PERIODONTAL DISEASE AND RISK OF ANKYLOSING SPONDYLITIS: A SYSTEMATIC REVIEW AND META-ANALYSISAnnals of the Rheumatic Diseases 2019;78:1829-1830. | Abstract |
| 162 | Fu W, Lv C, Zou L, Song F, Zeng X, Wang C, Yan S, Gan Y, Chen F, Lu Z, Cao S. Meta-analysis on the association between the frequency of tooth brushing and diabetes mellitus risk. Diabetes Metab Res Rev. 2019 Jul;35(5):e3141. doi: 10.1002/dmrr.3141. Epub 2019 Mar 18. PMID: 30758127. | Unrelated |
| 163 | Kellesarian SV, Yunker M, Malmstrom H, Almas K, Romanos GE, Javed F. Am J Mens Health. 2018 Nov;12(6):1976-1984. doi: 10.1177/1557988316655529. Epub 2016 Jun 23. | No control group |
| 164 | Acharya A, Khan S, Hoang H, Bettiol S, Goldberg L, Crocombe L. BMC Health Serv Res. 2018 Dec 3;18(1):921. doi: 10.1186/s12913-018-3733-2. | No control group |
| 165 | El Howati A, Tappuni A. J Investig Clin Dent. 2018 Nov;9(4):e12351. doi: 10.1111/jicd.12351. Epub 2018 Jul 17. | No Meta-Analysis |
| 166 | Pillai RS, Iyer K, Spin-Neto R, Kothari SF, Nielsen JF, Kothari M. Cerebrovasc Dis Extra. 2018;8(1):1-15. doi: 10.1159/000484989. Epub 2018 Jan 9. | No Meta-Analysis |
| 167 | Poudel P, Griffiths R, Wong VW, Arora A, Flack JR, Khoo CL, George A. BMC Public Health. 2018 May 2;18(1):577. doi: 10.1186/s12889-018-5485-7. | Unrelated |
| 168 | Delwel S, Binnekade TT, Perez RSGM, Hertogh CMPM, Scherder EJA, Lobbezoo F. Clin Oral Investig. 2018 Jan;22(1):93-108. doi: 10.1007/s00784-017-2264-2. Epub 2017 Nov 15. | No Meta-Analysis |
| 169 | Scalioni FAR, Carrada CF, Martins CC, Ribeiro RA, Paiva SM. J Am Dent Assoc. 2018 Jul;149(7):628-639.e11. doi: 10.1016/j.adaj.2018.03.010. Epub 2018 May 18. | No Meta-Analysis |
| 170 | Varela-López A, Navarro-Hortal MD, Giampieri F, Bullón P, Battino M, Quiles JL. Molecules. 2018 May 20;23(5):1226. doi: 10.3390/molecules23051226. | Unrelated |
| 171 | Kellesarian SV, Kellesarian TV, Ros Malignaggi V, Al-Askar M, Ghanem A, Malmstrom H, Javed F. Am J Mens Health. 2018 Mar;12(2):338-346. doi: 10.1177/1557988316639050. Epub 2016 Mar 29. | No Meta-Analysis |
| 172 | Najeeb S, Zafar MS, Khurshid Z, Zohaib S, Madathil SA, Mali M, Almas K. Saudi Pharm J. 2018 Jul;26(5):634-642. doi: 10.1016/j.jsps.2018.02.029. Epub 2018 Feb 16. | Included animal studies |
| 173 | Palla B, Burian E, Fliefel R, Otto S. Clin Oral Investig. 2018 Jan;22(1):1-27. doi: 10.1007/s00784-017-2124-0. Epub 2017 Jun 14. | No Meta-Analysis |
| 174 | Alakhali MS, Al-Maweri SA, Al-Shamiri HM, Al-Haddad K, Halboub E. Clin Oral Investig. 2018 Dec;22(9):2965-2974. doi: 10.1007/s00784-018-2726-1. Epub 2018 Oct 24. | No Meta-Analysis |
| 175 | Liu LS, Gkranias N, Farias B, Spratt D, Donos N. Clin Oral Investig. 2018 Nov;22(8):2743-2762. doi: 10.1007/s00784-018-2660-2. Epub 2018 Oct 10. | No Meta-Analysis |
| 176 | Pinto JPNS, Goergen J, Muniz FWMG, Haas AN. J Periodontal Res. 2018 Jun;53(3):298-305. doi: 10.1111/jre.12531. Epub 2018 Mar 1. | Unrelated |
| 177 | Almeida APCPSC, Fagundes NCF, Maia LC, Lima RR. Curr Vasc Pharmacol. 2018;16(6):569-582. doi: 10.2174/1570161115666170830141852. | No Meta-Analysis |
| 178 | Butala S, Palomo L. J Evid Based Dent Pract. 2018 Dec;18(4):352-354. doi: 10.1016/j.jebdp.2018.09.004. Epub 2018 Oct 4. | Commentary |
| 179 | Rangel-Rincón LJ, Vivares-Builes AM, Botero JE, Agudelo-Suárez AA. J Evid Based Dent Pract. 2018 Sep;18(3):218-239. doi: 10.1016/j.jebdp.2017.10.011. Epub 2017 Nov 4. | No Meta-Analysis |
| 180 | Khan S, Barrington G, Bettiol S, Barnett T, Crocombe L. Obes Rev. 2018 Jun;19(6):852-883. doi: 10.1111/obr.12668. Epub 2018 Jan 19. | No Meta-Analysis |
| 181 | Perić M, Cavalier E, Toma S, Lasserre JF. J Periodontal Res. 2018 Oct;53(5):645-656. doi: 10.1111/jre.12560. Epub 2018 Jun 2. | Unrelated |
| 182 | Graziani F, Gennai S, Solini A, Petrini M. J Clin Periodontol. 2018 Feb;45(2):167-187. doi: 10.1111/jcpe.12837. Epub 2017 Dec 26. | No Meta-Analysis |
| 183 | de Morais EF, Dantas AN, Pinheiro JC, Leite RB, Galvao Barboza CA, de Vasconcelos Gurgel BC, de Almeida Freitas R. Arch Oral Biol. 2018 Mar;87:43-51. doi: 10.1016/j.archoralbio.2017.12.008. Epub 2017 Dec 12. | No Meta-Analysis |
| 184 | Mergoni G, Percudani D, Lodi G, Bertani P, Manfredi M. J Endod. 2018 Nov;44(11):1616-1625.e9. doi: 10.1016/j.joen.2018.07.016. Epub 2018 Sep 18. | Unrelated |
| 185 | da Silva JC, Muniz FWMG, Oballe HJR, Andrades M, Rösing CK, Cavagni J. J Clin Periodontol. 2018 Oct;45(10):1222-1237. doi: 10.1111/jcpe.12993. Epub 2018 Aug 28. | No Meta-Analysis |
| 186 | Koka S, Gupta A. J Prosthodont Res. 2018 Apr;62(2):134-151. doi: 10.1016/j.jpor.2017.08.003. Epub 2017 Aug 30. | No Meta-Analysis |
| 187 | Ranjan R, Abhinay A, Mishra M. Neurol India. 2018 Mar-Apr;66(2):344-351. doi: 10.4103/0028-3886.227315. | No Meta-Analysis |
| 188 | Mashhadiabbas F, Neamatzadeh H, Nasiri R, Foroughi E, Farahnak S, Piroozmand P, Mazaheri M, Zare-Shehneh M. Dent Res J (Isfahan). 2018 May-Jun;15(3):155-165. | Unrelated |
| 189 | Martins OFM, Chaves Junior CM, Rossi RRP, Cunali PA, Dal-Fabbro C, Bittencourt L. Dental Press J Orthod. 2018 Aug 1;23(4):45-54. doi: 10.1590/2177-6709.23.4.045-054.oar. | No Meta-Analysis |
| 190 | Suvan JE, Finer N, D'Aiuto F. Periodontol 2000. 2018 Oct;78(1):98-128. doi: 10.1111/prd.12239. | Not a Systematic Review |
| 191 | Madianos PN, Koromantzos PA. J Clin Periodontol. 2018 Feb;45(2):188-195. doi: 10.1111/jcpe.12836. Epub 2017 Dec 26. | No Meta-Analysis |
| 192 | Martynowicz H, Smardz J, Wieczorek T, Mazur G, Poreba R, Skomro R, Zietek M, Wojakowska A, Michalek M, Wieckiewicz M. J Clin Med. 2018 Aug 23;7(9):233. doi: 10.3390/jcm7090233. | Unrelated |
| 193 | Kotsakis GA, Lian Q, Ioannou AL, Michalowicz BS, John MT, Chu H. J Periodontol. 2018 May;89(5):558-570. doi: 10.1002/JPER.17-0368. | Unrelated |
| 194 | Hashemi A, Bahrololoomi Z, Salarian S. Relationship Between Early Childhood Caries and Anemia: A Systematic Review. Iran J Ped Hematol Oncol. 2018; 8 (2) :126-138 | No Meta-Analysis |
| 195 | Sun D, Ye T, Ren P, Yu S. Prevalence and etiology of oral diseases in drug-addicted populations: a systematic review. Int J Clin Exp Med 2018;11(7):6521-6531 | No Meta-Analysis |
| 196 | Papi P, Letizia C, Pilloni A, Petramala L, Saracino V, Rosella D, Pompa G. Peri-implant diseases and metabolic syndrome components: a systematic review. Eur Rev Med Pharmacol Sci. 2018 Feb;22(4):866-875. doi: 10.26355/eurrev_201802_14364. PMID: 29509232. | No Meta-Analysis |
| 197 | Mac Giolla Phadraig C, Nunn J, McCallion P, McCarron M. Prevalence of edentulism among adults with intellectual disabilities: A narrative review informed by systematic review principles. Spec Care Dentist. 2018 Jul;38(4):191-200. doi: 10.1111/scd.12300. Epub 2018 Jun 8. PMID: 29882327. | No Meta-Analysis |
| 198 | Ungprasert P, Wijarnpreecha K, Cheungpasitporn W. Periodontal Disease Is Associated with an Increased Risk of Ankylosing Spondylitis: A Systematic Review and Meta-Analysis [abstract]. Arthritis Rheumatol. 2018; 70 (suppl 10). https://acrabstracts.org/abstract/periodontal-disease-is-associated-with-an-increased-risk-of-ankylosing-spondylitis-a-systematic-review-and-meta-analysis/. Accessed January 25, 2022. | Abstract |
| 199 | Brignardello-Petersen R. Human herpesviruses may be associated with the development of aggressive periodontitis. J Am Dent Assoc. 2018 Jul;149(7):e102. doi: 10.1016/j.adaj.2018.01.036. Epub 2018 Mar 13. PMID: 29548690. | Abstract |
| 200 | Delwel S, Binnekade TT, Perez RS, Hertogh CM, Scherder EJ, Lobbezoo F. Clin Oral Investig. 2017 Jan;21(1):17-32. doi: 10.1007/s00784-016-1934-9. Epub 2016 Sep 8. | No Meta-Analysis |
| 201 | da Silva SN, Gimenez T, Souza RC, Mello-Moura ACV, Raggio DP, Morimoto S, Lara JS, Soares GC, Tedesco TK. Int J Paediatr Dent. 2017 Sep;27(5):388-398. doi: 10.1111/ipd.12274. Epub 2016 Oct 31. | No control group |
| 202 | Chapple IL, Bouchard P, Cagetti MG, Campus G, Carra MC, Cocco F, Nibali L, Hujoel P, Laine ML, Lingstrom P, Manton DJ, Montero E, Pitts N, Rangé H, Schlueter N, Teughels W, Twetman S, Van Loveren C, Van der Weijden F, Vieira AR, Schulte AG. J Clin Periodontol. 2017 Mar;44 Suppl 18:S39-S51. doi: 10.1111/jcpe.12685. | Consensus |
| 203 | González Navarro B, Pintó Sala X, Jané Salas E. Med Clin (Barc). 2017 Sep 8;149(5):211-216. doi: 10.1016/j.medcli.2017.05.010. Epub 2017 Jun 20. | No Meta-Analysis |
| 204 | Foley NC, Affoo RH, Siqueira WL, Martin RE. JDR Clin Trans Res. 2017 Oct;2(4):330-342. doi: 10.1177/2380084417714789. Epub 2017 Jul 7. | No control group |
| 205 | Mauri-Obradors E, Estrugo-Devesa A, Jané-Salas E, Viñas M, López-López J. Med Oral Patol Oral Cir Bucal. 2017 Sep 1;22(5):e586-e594. doi: 10.4317/medoral.21655. | No control group |
| 206 | Pedrosa MS, de Paiva M, Oliveira L, Pereira S, da Silva C, Pompeu J. Aust Dent J. 2017 Dec;62(4):404-411. doi: 10.1111/adj.12516. Epub 2017 Jun 28. | No control group |
| 207 | Kapferer-Seebacher I, Lundberg P, Malfait F, Zschocke J. J Clin Periodontol. 2017 Nov;44(11):1088-1100. doi: 10.1111/jcpe.12807. Epub 2017 Sep 25. | No Meta-Analysis |
| 208 | Kothari M, Pillai RS, Kothari SF, Spin-Neto R, Kumar A, Nielsen JF. Oral Surg Oral Med Oral Pathol Oral Radiol. 2017 Feb;123(2):205-219.e7. doi: 10.1016/j.oooo.2016.10.024. Epub 2016 Nov 11. | No Meta-Analysis |
| 209 | Kellesarian SV, Malignaggi VR, Kellesarian TV, Al-Kheraif AA, Alwageet MM, Malmstrom H, Romanos GE, Javed F. Int J Impot Res. 2017 May;29(3):89-95. doi: 10.1038/ijir.2017.7. Epub 2017 Mar 9. | No Meta-Analysis |
| 210 | Hirsch V, Wolgin M, Mitronin AV, Kielbassa AM. Arch Oral Biol. 2017 Oct;82:38-46. doi: 10.1016/j.archoralbio.2017.05.008. Epub 2017 Jun 1. | Unrelated |
| 211 | Abduljabbar T, Javed F, Shah A, Samer MS, Vohra F, Akram Z. Lasers Med Sci. 2017 Feb;32(2):449-459. doi: 10.1007/s10103-016-2086-5. Epub 2016 Sep 29. | No Meta-Analysis |
| 212 | Al-Hamoudi N. Photodiagnosis Photodyn Ther. 2017 Sep;19:375-382. doi: 10.1016/j.pdpdt.2017.05.018. Epub 2017 May 27. | Unrelated |
| 213 | Li C, Lv Z, Shi Z, Zhu Y, Wu Y, Li L, Iheozor-Ejiofor Z. Cochrane Database Syst Rev. 2017 Nov 7;11(11):CD009197. doi: 10.1002/14651858.CD009197.pub3. | No Meta-Analysis |
| 214 | Goyal L, Goyal T, Gupta ND. J Midlife Health. 2017 Oct-Dec;8(4):151-158. doi: 10.4103/jmh.JMH_55_17. | Commentary |
| 215 | Martinez-Herrera M, Silvestre-Rangil J, Silvestre FJ. Med Oral Patol Oral Cir Bucal. 2017 Nov 1;22(6):e708-e715. doi: 10.4317/medoral.21786. | No Meta-Analysis |
| 216 | Berlin-Broner Y, Febbraio M, Levin L. Int Endod J. 2017 Sep;50(9):847-859. doi: 10.1111/iej.12710. Epub 2016 Nov 19. | No Meta-Analysis |
| 217 | Penoni DC, Fidalgo TK, Torres SR, Varela VM, Masterson D, Leão AT, Maia LC. J Dent Res. 2017 Mar;96(3):261-269. doi: 10.1177/0022034516682017. Epub 2017 Jan 3. | Unrelated |
| 218 | Hasuike A, Iguchi S, Suzuki D, Kawano E, Sato S. Med Oral Patol Oral Cir Bucal. 2017 Mar 1;22(2):e167-e176. doi: 10.4317/medoral.21555. | No Meta-Analysis |
| 219 | Cotti E, Arrica M, Di Lenarda A, Serri SB, Bassareo P, Padeletti L, Mercuro G. Eur J Prev Cardiol. 2017 Mar;24(4):409-425. doi: 10.1177/2047487316682348. Epub 2017 Jan 17. | No Meta-Analysis |
| 220 | Cheraghi Z, Doosti-Irani A. Int J Impot Res. 2017 Nov;29(6):262. doi: 10.1038/ijir.2017.33. | Letter to the editor |
| 221 | Kellesarian SV, Malmstrom H, Abduljabbar T, Vohra F, Kellesarian TV, Javed F, Romanos GE. Am J Mens Health. 2017 Mar;11(2):443-453. doi: 10.1177/1557988316667692. Epub 2016 Sep 21. | No Meta-Analysis |
| 222 | Bender P, Bürgin WB, Sculean A, Eick S. Clin Oral Investig. 2017 Jan;21(1):33-42. doi: 10.1007/s00784-016-1938-5. Epub 2016 Aug 25. | Unrelated |
| 223 | Bacevic M, Brkovic B, Albert A, Rompen E, Radermecker RP, Lambert F. Calcif Tissue Int. 2017 Dec;101(6):553-563. doi: 10.1007/s00223-017-0327-7. Epub 2017 Oct 24. | No Meta-Analysis |
| 224 | Tonsekar PP, Jiang SS, Yue G. Gerodontology. 2017 Jun;34(2):151-163. doi: 10.1111/ger.12261. Epub 2017 Feb 7. | No Meta-Analysis |
| 225 | Tettamanti L, Lauritano D, Nardone M, Gargari M, Silvestre-Rangil J, Gavoglio P, Tagliabue A. Oral Implantol (Rome). 2017 Sep 27;10(2):112-118. doi: 10.11138/orl/2017.10.2.112. eCollection 2017 Apr-Jun. | No Meta-Analysis |
| 226 | Qiu Q, Zhang F, Zhu W, Wu J, Liang M. Biol Trace Elem Res. 2017 May;177(1):53-63. doi: 10.1007/s12011-016-0877-y. Epub 2016 Oct 26. | Unrelated |
| 227 | Baghaie H, Kisely S, Forbes M, Sawyer E, Siskind DJ. A systematic review and meta-analysis of the association between poor oral health and substance abuse. Addiction. 2017 May;112(5):765-779. doi: 10.1111/add.13754. Epub 2017 Mar 16. PMID: 28299855. | Unrelated |
| 228 | Aminoshariae A, Kulild JC, Mickel A, Fouad AF. Association between Systemic Diseases and Endodontic Outcome: A Systematic Review. J Endod. 2017 Apr;43(4):514-519. doi: 10.1016/j.joen.2016.11.008. Epub 2017 Feb 9. PMID: 28190585. | No Meta-Analysis |
| 229 | Tada A, Miura H. Association between mastication and cognitive status: A systematic review. Arch Gerontol Geriatr. 2017 May-Jun;70:44-53. doi: 10.1016/j.archger.2016.12.006. Epub 2016 Dec 14. PMID: 28042986. | No Meta-Analysis |
| 230 | Gilheaney Ó, Zgaga L, Harpur I, Sheaf G, Kiefer L, Béchet S, Walshe M. The Prevalence of Oropharyngeal Dysphagia in Adults Presenting with Temporomandibular Disorders Associated with Rheumatoid Arthritis: A Systematic Review and Meta-analysis. Dysphagia. 2017 Oct;32(5):587-600. doi: 10.1007/s00455-017-9808-0. Epub 2017 May 16. PMID: 28508937. | No control group |
| 231 | Abu-Ashour W, Twells L, Valcour J, Randell A, Donnan J, Howse P, Gamble JM. The association between diabetes mellitus and incident infections: a systematic review and meta-analysis of observational studies. BMJ Open Diabetes Res Care. 2017 May 27;5(1):e000336. doi: 10.1136/bmjdrc-2016-000336. PMID: 28761647; PMCID: PMC5530269. | Unrelated |
| 232 | Kaye EK. Limited Evidence Suggests Tooth Loss is Associated With Increased Risk of Cognitive Impairment. J Evid Based Dent Pract. 2017 Mar;17(1):42-44. doi: 10.1016/j.jebdp.2017.01.006. Epub 2017 Jan 30. PMID: 28259313. | Commentary |
| 233 | Molina-García A, Castellanos-Cosano L, Machuca-Portillo G, Posada-de la Paz M. Med Oral Patol Oral Cir Bucal. 2016 Sep 1;21(5):e587-94. doi: 10.4317/medoral.20972. | No control group |
| 234 | Figueiredo Dde R, Bastos JL, Silva L, Peres KG. Community Dent Oral Epidemiol. 2016 Apr;44(2):180-7. doi: 10.1111/cdoe.12203. Epub 2015 Nov 25. | No Meta-Analysis |
| 235 | Diéguez-Pérez M, de Nova-García MJ, Mourelle-Martínez MR, Bartolomé-Villar B. J Clin Exp Dent. 2016 Jul 1;8(3):e337-43. doi: 10.4317/jced.52922. eCollection 2016 Jul. | No control group |
| 236 | Wu B, Fillenbaum GG, Plassman BL, Guo L. J Am Geriatr Soc. 2016 Apr;64(4):739-51. doi: 10.1111/jgs.14036. Epub 2016 Apr 1. | No control group |
| 237 | Varela-López A, Giampieri F, Bullón P, Battino M, Quiles JL. Int J Mol Sci. 2016 Jul 25;17(8):1202. doi: 10.3390/ijms17081202. | No control group |
| 238 | Almeida FT, Pachêco-Pereira C, Porporatti AL, Flores-Mir C, Leite AF, De Luca Canto G, Guerra EN. J Gastroenterol Hepatol. 2016 Mar;31(3):527-40. doi: 10.1111/jgh.13149. | No control group |
| 239 | Silva TD, Ferreira CB, Leite GB, de Menezes Pontes JR, Antunes HS. Ecancermedicalscience. 2016 Aug 17;10:665. doi: 10.3332/ecancer.2016.665. eCollection 2016. | No control group |
| 240 | Javed F, Malmstrom H, Kellesarian SV, Al-Kheraif AA, Vohra F, Romanos GE. Implant Dent. 2016 Apr;25(2):281-7. doi: 10.1097/ID.0000000000000390. | Unrelated |
| 241 | Okuno K, Pliska BT, Hamoda M, Lowe AA, Almeida FR. Sleep Med Rev. 2016 Dec;30:25-33. doi: 10.1016/j.smrv.2015.11.007. Epub 2015 Dec 8. | Unrelated |
| 242 | Wu B, Fillenbaum GG, Plassman BL, Guo L. J Am Geriatr Soc. 2016 Aug;64(8):1752. doi: 10.1111/jgs.14572. | No Meta-Analysis |
| 243 | Nascimento GG, Leite FR, Correa MB, Peres MA, Demarco FF. Clin Oral Investig. 2016 May;20(4):639-47. doi: 10.1007/s00784-015-1678-y. Epub 2015 Dec 1. | Unrelated |
| 244 | Rodríguez-Medina C, Agudelo-Suárez AA, Botero JE. J Evid Based Dent Pract. 2016 Dec;16(4):236-238. doi: 10.1016/j.jebdp.2016.11.005. Epub 2016 Nov 12. | Commentary |
| 245 | Javed F, Warnakulasuriya S. Crit Rev Oncol Hematol. 2016 Jan;97:197-205. doi: 10.1016/j.critrevonc.2015.08.018. Epub 2015 Aug 20. | No Meta-Analysis |
| 246 | Akram Z, Abduljabbar T, Sauro S, Daood U. Photodiagnosis Photodyn Ther. 2016 Dec;16:142-153. doi: 10.1016/j.pdpdt.2016.09.004. Epub 2016 Sep 9. | No Meta-Analysis |
| 247 | Annibali S, Pranno N, Cristalli MP, La Monaca G, Polimeni A. Implant Dent. 2016 Oct;25(5):663-74. doi: 10.1097/ID.0000000000000478. | Unrelated |
| 248 | Khalighinejad N, Aminoshariae MR, Aminoshariae A, Kulild JC, Mickel A, Fouad AF. J Endod. 2016 Oct;42(10):1427-34. doi: 10.1016/j.joen.2016.07.007. Epub 2016 Aug 31. | No Meta-Analysis |
| 249 | Botero JE, Rodríguez C, Agudelo-Suarez AA. Aust Dent J. 2016 Jun;61(2):134-48. doi: 10.1111/adj.12413. Epub 2016 Feb 26. | No Meta-Analysis |
| 250 | Jiang H, Xiong X. J Evid Based Dent Pract. 2016 Jun;16(2):121-3. doi: 10.1016/j.jebdp.2016.06.001. Epub 2016 Jun 4. | Commentary |
| 251 | Teshome A, Yitayeh A. Pan Afr Med J. 2016 Jul 12;24:215. doi: 10.11604/pamj.2016.24.215.8727. eCollection 2016. | No Meta-Analysis |
| 252 | Silvestre FJ, Silvestre-Rangil J, Bagán L, Bagán JV. Med Oral Patol Oral Cir Bucal. 2016 May 1;21(3):e349-54. doi: 10.4317/medoral.20974. | No Meta-Analysis |
| 253 | Mulimani P, Ballas SK, Abas AB, Karanth L. Cochrane Database Syst Rev. 2016 Apr 22;4:CD011633. doi: 10.1002/14651858.CD011633.pub2. | Unrelated |
| 254 | Rovai ES, Souto ML, Ganhito JA, Holzhausen M, Chambrone L, Pannuti CM. J Periodontol. 2016 Dec;87(12):1406-1417. doi: 10.1902/jop.2016.160214. Epub 2016 Jul 29. | No control group |
| 255 | Grellmann AP, Sfreddo CS, Maier J, Lenzi TL, Zanatta FB. J Clin Periodontol. 2016 Mar;43(3):250-60. doi: 10.1111/jcpe.12514. | No control group |
| 256 | Carvalho FR, Lentini-Oliveira DA, Prado LB, Prado GF, Carvalho LB. Cochrane Database Syst Rev. 2016 Oct 5;10(10):CD005520. doi: 10.1002/14651858.CD005520.pub3. | No Meta-Analysis |
| 257 | Pérez-Losada FL, Jané-Salas E, Sabater-Recolons MM, Estrugo-Devesa A, Segura-Egea JJ, López-López J. Med Oral Patol Oral Cir Bucal. 2016 Jul 1;21(4):e440-6. doi: 10.4317/medoral.21048. | No Meta-Analysis |
| 258 | Cagnani A, Barros AMS, Sousa LLA, Zanin L, Bergamaschi CC, Peruzzo DC, and Flório FM. 2016. Periodontal disease as a risk factor for aspiration pneumonia: a systematic review . Bioscience Journal, vol. 32:3. DOI 10.14393/BJ-v32n3a2016-33210. | No Meta-Analysis |
| 259 | Fuggle N, Smith T, Kaul A, et alFRI0111 Dental Association or Incidental Finding? A Meta-Analysis and Systematic Review of The Relationship between Rheumatoid Arthritis and PeriodontitisAnnals of the Rheumatic Diseases 2016;75:468. | Abstract |
| 260 | López-Pintor RM, Casañas E, González-Serrano J, Serrano J, Ramírez L, de Arriba L, Hernández G. Xerostomia, Hyposalivation, and Salivary Flow in Diabetes Patients. J Diabetes Res. 2016;2016:4372852. doi: 10.1155/2016/4372852. Epub 2016 Jul 10. PMID: 27478847; PMCID: PMC4958434. | No Meta-Analysis |
| 261 | Ashley P, Di Iorio A, Cole E, Tanday A, Needleman I. Br J Sports Med. 2015 Jan;49(1):14-9. doi: 10.1136/bjsports-2014-093617. Epub 2014 Nov 11. | Unrelated |
| 262 | Ismail AF, McGrath CP, Yiu CK. Diabetes Res Clin Pract. 2015 Jun;108(3):369-81. doi: 10.1016/j.diabres.2015.03.003. Epub 2015 Mar 14. | No Meta-Analysis |
| 263 | Dai R, Lam OL, Lo EC, Li LS, Wen Y, McGrath C. J Dent. 2015 Feb;43(2):171-80. doi: 10.1016/j.jdent.2014.06.005. Epub 2014 Jun 21. | No Meta-Analysis |
| 264 | Moazzam AA, Rajagopal SM, Sedghizadeh PP, Zada G, Habibian M. J Clin Neurosci. 2015 May;22(5):800-6. doi: 10.1016/j.jocn.2014.11.015. Epub 2015 Mar 21. | No control group |
| 265 | Grønkjær LL. SAGE Open Med. 2015 Sep 9;3:2050312115601122. doi: 10.1177/2050312115601122. eCollection 2015. | No Meta-Analysis |
| 266 | El-Rabbany M, Zaghlol N, Bhandari M, Azarpazhooh A. Int J Nurs Stud. 2015 Jan;52(1):452-64. doi: 10.1016/j.ijnurstu.2014.07.010. Epub 2014 Jul 27. | No Meta-Analysis |
| 267 | Keller A, Rohde JF, Raymond K, Heitmann BL. J Periodontol. 2015 Jun;86(6):766-76. doi: 10.1902/jop.2015.140589. Epub 2015 Feb 12. | No Meta-Analysis |
| 268 | Mendes L, Azevedo NF, Felino A, Pinto MG. Virulence. 2015;6(3):208-15. doi: 10.4161/21505594.2014.984566. | No Meta-Analysis |
| 269 | López NJ, Uribe S, Martinez B. Periodontol 2000. 2015 Feb;67(1):87-130. doi: 10.1111/prd.12073. | No Meta-Analysis |
| 270 | Cortela DC, de Souza Junior AL, Virmond MC, Ignotti E. Mediators Inflamm. 2015;2015:548540. doi: 10.1155/2015/548540. Epub 2015 Aug 3. | No Meta-Analysis |
| 271 | Mauri-Obradors E, Jané-Salas E, Sabater-Recolons Mdel M, Vinas M, López-López J. Odontology. 2015 Sep;103(3):301-13. doi: 10.1007/s10266-014-0165-2. Epub 2014 Jul 26. | No Meta-Analysis |
| 272 | Schwendicke F, Karimbux N, Allareddy V, Gluud C. PLoS One. 2015 Jun 2;10(6):e0129060. doi: 10.1371/journal.pone.0129060. eCollection 2015. | Data from a previous SR |
| 273 | Estanislau IM, Terceiro IR, Lisboa MR, Teles Pde B, Carvalho Rde S, Martins RS, Moreira MM. Br J Clin Pharmacol. 2015 Jun;79(6):877-85. doi: 10.1111/bcp.12564. | No Meta-Analysis |
| 274 | Amrutiya MR, Deshpande N. Role of Obesity in Chronic Periodontal Diseases: A Systematic Review. 2015 J Dent & Oral Disord. 2016; 2(2): 1012. | No Meta-Analysis |
| 275 | Lieshout HF, Bots CP. Clin Oral Investig. 2014 Jan;18(1):17-24. doi: 10.1007/s00784-013-1034-z. Epub 2013 Jul 20. | No control group |
| 276 | Genco RJ, Genco FD. J Evid Based Dent Pract. 2014 Jun;14 Suppl:4-16. doi: 10.1016/j.jebdp.2014.03.003. Epub 2014 Mar 27. | No Meta-Analysis |
| 277 | Ruospo M, Palmer SC, Craig JC, Gentile G, Johnson DW, Ford PJ, Tonelli M, Petruzzi M, De Benedittis M, Strippoli GF. Nephrol Dial Transplant. 2014 Feb;29(2):364-75. doi: 10.1093/ndt/gft401. Epub 2013 Sep 29. | No Meta-Analysis |
| 278 | Tada A, Miura H. Arch Gerontol Geriatr. 2014 Nov-Dec;59(3):497-505. doi: 10.1016/j.archger.2014.08.005. Epub 2014 Aug 17. | No Meta-Analysis |
| 279 | Kassebaum NJ, Bernabé E, Dahiya M, Bhandari B, Murray CJ, Marcenes W. J Dent Res. 2014 Nov;93(11):1045-53. doi: 10.1177/0022034514552491. Epub 2014 Sep 26. | Unrelated |
| 280 | Pushparani DS. Curr Diabetes Rev. 2014;10(6):397-401. doi: 10.2174/1573399810666141121161514. | No Meta-Analysis |
| 281 | Hurst D. Evid Based Dent. 2014 Dec;15(4):102-3. doi: 10.1038/sj.ebd.6401057. | Commentary |
| 282 | Li C, Lv Z, Shi Z, Zhu Y, Wu Y, Li L, Iheozor-Ejiofor Z. Cochrane Database Syst Rev. 2014 Aug 15;(8):CD009197. doi: 10.1002/14651858.CD009197.pub2. | No Meta-Analysis |
| 283 | Horliana AC, Chambrone L, Foz AM, Artese HP, Rabelo Mde S, Pannuti CM, Romito GA. PLoS One. 2014 May 28;9(5):e98271. doi: 10.1371/journal.pone.0098271. eCollection 2014. | No Meta-Analysis |
| 284 | Kulkarni V, Bhatavadekar NB, Uttamani JR. J Calif Dent Assoc. 2014 May;42(5):302-11. | No Meta-Analysis |
| 285 | Awan KH, Patil S, Habib SR, Pejcic A, Zain RB. J Contemp Dent Pract. 2014 Nov 1;15(6):812-7. doi: 10.5005/jp-journals-10024-1623. | No Meta-Analysis |
| 286 | Lassi ZS, Imam AM, Dean SV, Bhutta ZA. Reprod Health. 2014 Sep 26;11 Suppl 3(Suppl 3):S4. doi: 10.1186/1742-4755-11-S3-S4. Epub 2014 Sep 26. | Unrelated |
| 287 | Lechien JR, Filleul O, Costa de Araujo P, Hsieh JW, Chantrain G, Saussez S. Int J Otolaryngol. 2014;2014:465173. doi: 10.1155/2014/465173. Epub 2014 Apr 8. | Unrelated |
| 288 | Al Sayed A, Anand PS, Kamath KP, Patil S, Preethanath RS, Anil S. ISRN Gastroenterol. 2014 Feb 20;2014:261369. doi: 10.1155/2014/261369. eCollection 2014. | No Meta-Analysis |
| 289 | Kassebaum NJ, Bertozzi-Villa A, Coggeshall MS, Shackelford KA, Steiner C, Heuton KR, Gonzalez-Medina D, Barber R, Huynh C, Dicker D, Templin T, Wolock TM, Ozgoren AA, Abd-Allah F, Abera SF, Abubakar I, Achoki T, Adelekan A, Ademi Z, Adou AK, Adsuar JC, Agardh EE, Akena D, Alasfoor D, Alemu ZA, Alfonso-Cristancho R, Alhabib S, Ali R, Al Kahbouri MJ, Alla F, Allen PJ, AlMazroa MA, Alsharif U, Alvarez E, Alvis-Guzmán N, Amankwaa AA, Amare AT, Amini H, Ammar W, Antonio CA, Anwari P, Arnlöv J, Arsenijevic VS, Artaman A, Asad MM, Asghar RJ, Assadi R, Atkins LS, Badawi A, Balakrishnan K, Basu A, Basu S, Beardsley J, Bedi N, Bekele T, Bell ML, Bernabe E, Beyene TJ, Bhutta Z, Bin Abdulhak A, Blore JD, Basara BB, Bose D, Breitborde N, Cárdenas R, Castañeda-Orjuela CA, Castro RE, Catalá-López F, Cavlin A, Chang JC, Che X, Christophi CA, Chugh SS, Cirillo M, Colquhoun SM, Cooper LT, Cooper C, da Costa Leite I, Dandona L, Dandona R, Davis A, Dayama A, Degenhardt L, De Leo D, del Pozo-Cruz B, Deribe K, Dessalegn M, deVeber GA, Dharmaratne SD, Dilmen U, Ding EL, Dorrington RE, Driscoll TR, Ermakov SP, Esteghamati A, Faraon EJ, Farzadfar F, Felicio MM, Fereshtehnejad SM, de Lima GM, et al. Lancet. 2014 Sep 13;384(9947):980-1004. doi: 10.1016/S0140-6736(14)60696-6. Epub 2014 May 2. | Unrelated |
| 290 | Andrade, MRTC; Antunes, LAA; Soares, RMDA; Leao, ATT; Maia, LC; Primo, LG. 2014. Lower dental caries prevalence associated to chronic kidney disease: a systematic review (vol 29, pg 467, 2014) | No Meta-Analysis |
| 291 | Clementini M, Rossetti PH, Penarrocha D, Micarelli C, Bonachela WC, Canullo L. Systemic risk factors for peri-implant bone loss: a systematic review and meta-analysis. Int J Oral Maxillofac Surg. 2014 Mar;43(3):323-34. doi: 10.1016/j.ijom.2013.11.012. Epub 2013 Dec 25. PMID: 24373525. | Unrelated |
| 292 | Hujoel PP. Nutr Rev. 2013 Feb;71(2):88-97. doi: 10.1111/j.1753-4887.2012.00544.x. Epub 2012 Nov 9. | Unrelated |
| 293 | van der Maarel-Wierink CD, Vanobbergen JN, Bronkhorst EM, Schols JM, de Baat C. Gerodontology. 2013 Mar;30(1):3-9. doi: 10.1111/j.1741-2358.2012.00637.x. Epub 2012 Mar 6. | No Meta-Analysis |
| 294 | Eliyas S, Al-Khayatt A, Porter RW, Briggs P. Cochrane Database Syst Rev. 2013 Feb 28;(2):CD008857. doi: 10.1002/14651858.CD008857.pub2. | No Meta-Analysis |
| 295 | Stadelmann P, Alessandri R, Eick S, Salvi GE, Surbek D, Sculean A. Clin Oral Investig. 2013 Jul;17(6):1453-63. doi: 10.1007/s00784-013-0952-0. Epub 2013 Mar 7. | No Meta-Analysis |
| 296 | Borgnakke WS. J Evid Based Dent Pract. 2013 Sep;13(3):88-90. doi: 10.1016/j.jebdp.2013.07.002. | Commentary |
| 297 | Kaur S, White S, Bartold PM. J Dent Res. 2013 May;92(5):399-408. doi: 10.1177/0022034513483142. Epub 2013 Mar 22. | No Meta-Analysis |
| 298 | Boillot A, Zoungas S, Mitchell P, Klein R, Klein B, Ikram MK, Klaver C, Wang JJ, Gopinath B, Tai ES, Neubauer AS, Hercberg S, Brazionis L, Saw SM, Wong TY, Czernichow S; META-EYE Study Group. PLoS One. 2013;8(2):e52708. doi: 10.1371/journal.pone.0052708. Epub 2013 Feb 6. | Unrelated |
| 299 | Ide M, Papapanou PN. J Periodontol. 2013 Apr;84(4 Suppl):S181-94. doi: 10.1902/jop.2013.134009. | No Meta-Analysis |
| 300 | Ide M, Papapanou PN. J Clin Periodontol. 2013 Apr;40 Suppl 14:S181-94. doi: 10.1111/jcpe.12063. | No Meta-Analysis |
| 301 | Borgnakke WS, Ylöstalo PV, Taylor GW, Genco RJ. J Clin Periodontol. 2013 Apr;40 Suppl 14:S135-52. doi: 10.1111/jcpe.12080. | No Meta-Analysis |
| 302 | Shah M, Muley A, Muley P. J Matern Fetal Neonatal Med. 2013 Nov;26(17):1691-5. doi: 10.3109/14767058.2013.799662. Epub 2013 May 23. | No Meta-Analysis |
| 303 | Kelly JT, Avila-Ortiz G, Allareddy V, Johnson GK, Elangovan S. J Am Dent Assoc. 2013 Apr;144(4):371-9. doi: 10.14219/jada.archive.2013.0130. | No Meta-Analysis |
| 304 | Kim Y, Nowzari H, Rich SK. Clin Implant Dent Relat Res. 2013 Oct;15(5):645-53. doi: 10.1111/j.1708-8208.2011.00407.x. Epub 2011 Dec 15. | No Meta-Analysis |
| 305 | Dietrich T, Sharma P, Walter C, Weston P, Beck J. J Periodontol. 2013 Apr;84(4 Suppl):S70-84. doi: 10.1902/jop.2013.134008. | No Meta-Analysis |
| 306 | Niederman R. Evid Based Dent. 2013 Dec;14(4):107-8. doi: 10.1038/sj.ebd.6400966. | Commentary |
| 307 | Marsicano JA, de Moura-Grec PG, Bonato RC, Sales-Peres Mde C, Sales-Peres A, Sales-Peres SH. Gastroesophageal reflux, dental erosion, and halitosis in epidemiological surveys: a systematic review. Eur J Gastroenterol Hepatol. 2013 Feb;25(2):135-41. doi: 10.1097/MEG.0b013e32835ae8f7. PMID: 23111415. | No Meta-Analysis |
| 308 | Mendz GL, Kaakoush NO, Quinlivan JA. Bacterial aetiological agents of intra-amniotic infections and preterm birth in pregnant women. Front Cell Infect Microbiol. 2013 Oct 16;3:58. doi: 10.3389/fcimb.2013.00058. PMID: 24137568; PMCID: PMC3797391. | Not a Systematic Review |
| 309 | Chestnutt IG. J Evid Based Dent Pract. 2012 Mar;12(1):26-7. doi: 10.1016/j.jebdp.2011.12.015. | Commentary |
| 310 | Shanthi V, Vanka A, Bhambal A, Saxena V, Saxena S, Kumar SS. Dent Res J (Isfahan). 2012 Jul;9(4):368-80. | No Meta-Analysis |
| 311 | Polzer I, Schwahn C, Völzke H, Mundt T, Biffar R. Clin Oral Investig. 2012 Apr;16(2):333-51. doi: 10.1007/s00784-011-0625-9. Epub 2011 Nov 17. | Failed to provide heterogeneity values |
| 312 | Hultin M, Davidson T, Gynther G, Helgesson G, Jemt T, Lekholm U, Nilner K, Nordenram G, Norlund A, Rohlin M, Sunnegardh-Gronberg K, Tranaeus S. Int J Prosthodont. 2012 Nov-Dec;25(6):543-52. | Unrelated |
| 313 | Freitas CO, Gomes-Filho IS, Naves RC, Nogueira Filho Gda R, Cruz SS, Santos CA, Dunningham L, Miranda LF, Barbosa MD. J Appl Oral Sci. 2012 Feb;20(1):1-8. doi: 10.1590/s1678-77572012000100002. | Failed to provide MA estimates |
| 314 | Stefler D, Bhopal R, Fischbacher CM. Public Health. 2012 May;126(5):397-409. doi: 10.1016/j.puhe.2012.01.033. Epub 2012 Apr 5. | No Meta-Analysis |
| 315 | Han JY, Reynolds MA. J Periodontal Implant Sci. 2012 Feb;42(1):3-12. doi: 10.5051/jpis.2012.42.1.3. Epub 2012 Feb 29. | Failed to provide MA estimates |
| 316 | Brocklehurst P, Tickle M, Glenny AM, Lewis MA, Pemberton MN, Taylor J, Walsh T, Riley P, Yates JM. Cochrane Database Syst Rev. 2012 Sep 12;(9):CD005411. doi: 10.1002/14651858.CD005411.pub2. | Unrelated |
| 317 | Chen LL, Li H, Zhang PP, Wang SM. J Periodontol. 2012 Sep;83(9):1095-103. doi: 10.1902/jop.2011.110518. Epub 2011 Dec 19. | Unrelated |
| 318 | Kaur S, White S, Bartold M. JBI Libr Syst Rev. 2012;10(42 Suppl):1-12. doi: 10.11124/jbisrir-2012-288. | No Meta-Analysis |
| 319 | Matthews D. Evid Based Dent. 2012;13(3):80. doi: 10.1038/sj.ebd.6400875. | Commentary |
| 320 | Xiao-Ping W, Wei L, Yu-Shan H. CORRELATION BETWEEN CHRONIC PERIODONTITIS AND CARDIOVASCULAR DISEASES: A SYSTEMATIC REVIEWHeart 2012;98:E93-E94. | Abstract |
| 321 | Terezakis E, Needleman I, Kumar N, Moles D, Agudo E. J Clin Periodontol. 2011 Jul;38(7):628-36. doi: 10.1111/j.1600-051X.2011.01727.x. Epub 2011 Apr 7. | No control group |
| 322 | Cornacchio AL, Burneo JG, Aragon CE. J Can Dent Assoc. 2011;77:b140. | No Meta-Analysis |
| 323 | van der Maarel-Wierink CD, Vanobbergen JN, Bronkhorst EM, Schols JM, de Baat C. J Am Med Dir Assoc. 2011 Jun;12(5):344-54. doi: 10.1016/j.jamda.2010.12.099. Epub 2011 Mar 21. | No Meta-Analysis |
| 324 | Fuertes-González MC, Silvestre FJ, Almerich-Silla JM. Med Oral Patol Oral Cir Bucal. 2011 Jan 1;16(1):e37-41. doi: 10.4317/medoral.16.e37. | No Meta-Analysis |
| 325 | Khokhar WA, Clifton A, Jones H, Tosh G. Cochrane Database Syst Rev. 2011 Nov 9;(11):CD008802. doi: 10.1002/14651858.CD008802.pub2. | No Meta-Analysis |
| 326 | Katz J, Bimstein E. Quintessence Int. 2011 Jul-Aug;42(7):595-9. | No Meta-Analysis |
| 327 | Preshaw PM, Taylor JJ. J Clin Periodontol. 2011 Mar;38 Suppl 11:60-84. doi: 10.1111/j.1600-051X.2010.01671.x. | No Meta-Analysis |
| 328 | Baccaglini L. J Am Dent Assoc. 2011 Oct;142(10):1192-3. doi: 10.14219/jada.archive.2011.0089. | Commentary |
| 329 | Sadighi Shamami M, Sadighi Shamami M, Amini S. Iran J Cancer Prev. 2011 Fall;4(4):189-98. | Not a Systematic Review |
| 330 | Anders PL, Davis EL. Spec Care Dentist. 2010 May-Jun;30(3):110-7. doi: 10.1111/j.1754-4505.2010.00136.x. | No Meta-Analysis |
| 331 | Hong CH, Napeñas JJ, Hodgson BD, Stokman MA, Mathers-Stauffer V, Elting LS, Spijkervet FK, Brennan MT; Dental Disease Section, Oral Care Study Group, Multi-national Association of Supportive Care in Cancer (MASCC)/International Society of Oral Oncology (ISOO). Support Care Cancer. 2010 Aug;18(8):1007-21. doi: 10.1007/s00520-010-0873-2. Epub 2010 May 7. | No Meta-Analysis |
| 332 | Worthington HV, Clarkson JE, Khalid T, Meyer S, McCabe M. Cochrane Database Syst Rev. 2010 Jul 7;2010(7):CD001972. doi: 10.1002/14651858.CD001972.pub4. | Unrelated |
| 333 | Martínez-Maestre MÁ, González-Cejudo C, Machuca G, Torrejón R, Castelo-Branco C. Climacteric. 2010 Dec;13(6):523-9. doi: 10.3109/13697137.2010.500749. Epub 2010 Aug 7. | No Meta-Analysis |
| 334 | Dhadse P, Gattani D, Mishra R. J Indian Soc Periodontol. 2010 Jul;14(3):148-54. doi: 10.4103/0972-124X.75908. | No Meta-Analysis |
| 335 | Pimentel Lopes De Oliveira GJ, Amaral Fontanari L, Chaves De Souza JA, Ribeiro Costa M, Cirelli JA. Minerva Stomatol. 2010 Oct;59(10):543-50. | No Meta-Analysis |
| 336 | Lalla RV, Latortue MC, Hong CH, Ariyawardana A, D'Amato-Palumbo S, Fischer DJ, Martof A, Nicolatou-Galitis O, Patton LL, Elting LS, Spijkervet FK, Brennan MT; Fungal Infections Section, Oral Care Study Group, Multinational Association of Supportive Care in Cancer (MASCC)/International Society of Oral Oncology (ISOO). A systematic review of oral fungal infections in patients receiving cancer therapy. Support Care Cancer. 2010 Aug;18(8):985-92. doi: 10.1007/s00520-010-0892-z. Epub 2010 May 8. PMID: 20449755; PMCID: PMC2914797. | No Meta-Analysis |
| 337 | Jensen SB, Pedersen AM, Vissink A, Andersen E, Brown CG, Davies AN, Dutilh J, Fulton JS, Jankovic L, Lopes NN, Mello AL, Muniz LV, Murdoch-Kinch CA, Nair RG, Napeñas JJ, Nogueira-Rodrigues A, Saunders D, Stirling B, von Bültzingslöwen I, Weikel DS, Elting LS, Spijkervet FK, Brennan MT; Salivary Gland Hypofunction/Xerostomia Section; Oral Care Study Group; Multinational Association of Supportive Care in Cancer (MASCC)/International Society of Oral Oncology (ISOO). A systematic review of salivary gland hypofunction and xerostomia induced by cancer therapies: management strategies and economic impact. Support Care Cancer. 2010 Aug;18(8):1061-79. doi: 10.1007/s00520-010-0837-6. Epub 2010 Mar 25. PMID: 20333412. | No Meta-Analysis |
| 338 | Jensen SB, Pedersen AM, Vissink A, Andersen E, Brown CG, Davies AN, Dutilh J, Fulton JS, Jankovic L, Lopes NN, Mello AL, Muniz LV, Murdoch-Kinch CA, Nair RG, Napeñas JJ, Nogueira-Rodrigues A, Saunders D, Stirling B, von Bültzingslöwen I, Weikel DS, Elting LS, Spijkervet FK, Brennan MT; Salivary Gland Hypofunction/Xerostomia Section, Oral Care Study Group, Multinational Association of Supportive Care in Cancer (MASCC)/International Society of Oral Oncology (ISOO). A systematic review of salivary gland hypofunction and xerostomia induced by cancer therapies: prevalence, severity and impact on quality of life. Support Care Cancer. 2010 Aug;18(8):1039-60. doi: 10.1007/s00520-010-0827-8. Epub 2010 Mar 17. PMID: 20237805. | No Meta-Analysis |
| 339 | McCracken G. Evid Based Dent. 2009;10(2):42. doi: 10.1038/sj.ebd.6400645. | Commentary |
| 340 | van der Putten GJ, Vanobbergen J, De Visschere L, Schols J, de Baat C. Nutrition. 2009 Jul-Aug;25(7-8):717-22. doi: 10.1016/j.nut.2009.01.012. | No Meta-Analysis |
| 341 | Javed F, Romanos GE. J Periodontol. 2009 Nov;80(11):1719-30. doi: 10.1902/jop.2009.090283. | No Meta-Analysis |
| 342 | Menezes EV, Yakoob MY, Soomro T, Haws RA, Darmstadt GL, Bhutta ZA. BMC Pregnancy Childbirth. 2009 May 7;9 Suppl 1(Suppl 1):S4. doi: 10.1186/1471-2393-9-S1-S4. | No Meta-Analysis |
| 343 | Garcia R. Evid Based Dent. 2009;10(1):20-1. doi: 10.1038/sj.ebd.6400633. | Commentary |
| 344 | Helfand M, Buckley D, Fleming C, Fu R, Freeman M, Humphrey L, Rogers K, Walker M. Rockville (MD): Agency for Healthcare Research and Quality (US); 2009 Oct. Report No.: 10-05141-EF-1. | No Meta-Analysis |
| 345 | Conde-Agudelo, A., & Romero, R. (2009). 794: Maternal periodontal disease and risk of preeclampsia: a systematic review and meta-analysis. American Journal of Obstetrics and Gynecology, 201(6), S285. doi:10.1016/j.ajog.2009.10.811 | Abstract |
| 346 | Chavarry, NGM; Vettore, MV; Sansone, C; Sheiham, A (2009). The Relationship Between Diabetes Mellitus and Destructive Periodontal Disease: A Meta-Analysis. Oral Health and Preventive Dentistry, 2:2009:107-127 | No Meta-Analysis |
| 347 | Vergnes JN. Evid Based Dent. 2008;9(2):46-7. doi: 10.1038/sj.ebd.6400580. | Commentary |
| 348 | Humphrey LL, Fu R, Buckley DI, Freeman M, Helfand M. J Gen Intern Med. 2008 Dec;23(12):2079-86. doi: 10.1007/s11606-008-0787-6. Epub 2008 Sep 20. | Failed to provide heterogeneity values |
| 349 | Rustveld LO, Kelsey SF, Sharma R. Association between maternal infections and preeclampsia: a systematic review of epidemiologic studies. Matern Child Health J. 2008 Mar;12(2):223-42. doi: 10.1007/s10995-007-0224-1. Epub 2007 Jun 19. PMID: 17577649. | Unrelated |
| 350 | Boren SA, Gunlock TL, Schaefer J, Albright A. Diabetes Educ. 2007 Nov-Dec;33(6):1053-77; discussion 1078-9. doi: 10.1177/0145721707309809. | No Meta-Analysis |
| 351 | Clarkson JE, Worthington HV, Eden OB. Cochrane Database Syst Rev. 2007 Jan 24;2007(1):CD003807. doi: 10.1002/14651858.CD003807.pub3. | Unrelated |
| 352 | Klokkevold PR, Han TJ. Int J Oral Maxillofac Implants. 2007;22 Suppl:173-202. | No Meta-Analysis |
| 353 | Alves C, Andion J, Brandão M, Menezes R. Arq Bras Endocrinol Metabol. 2007 Oct;51(7):1050-7. doi: 10.1590/s0004-27302007000700005. | No Meta-Analysis |
| 354 | Worthington HV, Clarkson JE, Eden OB. Cochrane Database Syst Rev. 2007 Apr 18;(2):CD001972. doi: 10.1002/14651858.CD001972.pub3. | Unrelated |
| 355 | Mustapha IZ, Debrey S, Oladubu M, Ugarte R. J Periodontol. 2007 Dec;78(12):2289-302. doi: 10.1902/jop.2007.070140. | Unrelated |
| 356 | Xiong X, Buekens P, Vastardis S, Yu SM. Obstet Gynecol Surv. 2007 Sep;62(9):605-15. doi: 10.1097/01.ogx.0000279292.63435.40. | No Meta-Analysis |
| 357 | Azarpazhooh A, Leake JL. J Periodontol. 2006 Sep;77(9):1465-82. doi: 10.1902/jop.2006.060010. | No Meta-Analysis |
| 358 | Georg D. Int J Evid Based Healthc. 2006 Mar;4(1):54-61. doi: 10.1111/j.1479-6988.2006.00032.x. | No Meta-Analysis |
| 359 | Vettore MV, Lamarca Gde A, Leão AT, Thomaz FB, Sheiham A, Leal Mdo C. Cad Saude Publica. 2006 Oct;22(10):2041-53. doi: 10.1590/s0102-311x2006001000010. | No Meta-Analysis |
| 360 | Nugent JL, Baker PN. BJOG. 2006 Jul;113(7):848; author reply 848-9. doi: 10.1111/j.1471-0528.2006.00969.x. Epub 2006 Jun 2. | Commentary |
| 361 | Xiong X, Buekens P, Fraser WD, Beck J, Offenbacher S. BJOG. 2006 Feb;113(2):135-43. doi: 10.1111/j.1471-0528.2005.00827.x. | No Meta-Analysis |
| 362 | Worthington HV, Eden OB, Clarkson JE. Cochrane Database Syst Rev. 2004 Oct 18;(4):CD003807. doi: 10.1002/14651858.CD003807.pub2. | Unrelated |
| 363 | Clarkson JE, Worthington HV, Eden OB. Cochrane Database Syst Rev. 2004;(1):CD001972. doi: 10.1002/14651858.CD001972.pub2. | Unrelated |
| 364 | Paquette DW. Compend Contin Educ Dent. 2004 Sep;25(9):681-2, 685-92; quiz 694. | No Meta-Analysis |
| 365 | Scannapieco FA, Bush RB, Paju S. Ann Periodontol. 2003 Dec;8(1):54-69. doi: 10.1902/annals.2003.8.1.54. | Unrelated |
| 366 | Scannapieco FA, Bush RB, Paju S. Ann Periodontol. 2003 Dec;8(1):38-53. doi: 10.1902/annals.2003.8.1.38. | No Meta-Analysis |
| 367 | Scannapieco FA, Bush RB, Paju S. Ann Periodontol. 2003 Dec;8(1):70-8. doi: 10.1902/annals.2003.8.1.70. | No Meta-Analysis |
| 368 | Ryan ME, Carnu O, Kamer A. J Am Dent Assoc. 2003 Oct;134 Spec No:34S-40S. doi: 10.14219/jada.archive.2003.0370. | No Meta-Analysis |
| 369 | Worthington HV, Clarkson JE. J Dent Educ. 2002 Aug;66(8):903-11. | Unrelated |
| 370 | Albougy HA, Naidoo S. SADJ. 2002 Dec;57(11):457-66. | Unrelated |
| 371 | Worthington HV, Clarkson JE, Eden OB. Cochrane Database Syst Rev. 2002;(3):CD003807. doi: 10.1002/14651858.CD003807. | No Meta-Analysis |
| 372 | Clarkson JE, Worthington HV, Eden OB. Cochrane Database Syst Rev. 2002;(1):CD001972. doi: 10.1002/14651858.CD001972. | Unrelated |
| 373 | Madianos PN, Bobetsis GA, Kinane DF. J Clin Periodontol. 2002;29 Suppl 3:22-36; discussion 37-8. doi: 10.1034/j.1600-051x.29.s3.2.x. | No Meta-Analysis |
| 374 | Patton LL, Shugars DA, Bonito AJ. J Am Dent Assoc. 2002 Feb;133(2):195-203. doi: 10.14219/jada.archive.2002.0144. | Unrelated |
| 375 | Garcia RI, Nunn ME, Vokonas PS. Ann Periodontol. 2001 Dec;6(1):71-7. doi: 10.1902/annals.2001.6.1.71. | No Meta-Analysis |
| 376 | Clarkson JE, Worthington HV, Eden OB. Cochrane Database Syst Rev. 2000;(2):CD000978. doi: 10.1002/14651858.CD000978. | Unrelated |
| 377 | Danesh J, Appleby P. Expert Opin Investig Drugs. 1998 May;7(5):691-713. doi: 10.1517/13543784.7.5.691. | Unrelated |
| 378 | Meurman JH, Pyrhönen S, Teerenhovi L, Lindqvist C. Oral Oncol. 1997 Nov;33(6):389-97. doi: 10.1016/s0964-1955(97)00032-8. | Not a Systematic Review |
