## Supplementary material for "An umbrella review of the evidence linking oral health and systemic health: from the prevalence to clinical and circulating markers": Supplementary data 3.docx

**Supplementary Data 3.** Summary of the country origin of the systematic review.

| n | Country | References |
| --- | --- | --- |
| 43 | Brazil | de Lima et al. (2020); Cademartori et al. (2018); Cerutti-Kopplin et al. (2016); Nascimento et al. (2015); Jerônimo et al. (2020); Galdino et al. (2021); Nascimento et al. (2016); Araújo et al. (2016); Ferreira et al. (2019); Ferreira et al. (2019); da Silva et al. (2021); Daudt et al. (2018); Moraschini et al. (2016); Moraschini et al. (2018); Moura-Grec et al. (2014); de Oliveira Ferreira et al. (2019); Esteves Lima et al. (2016); Rosa et al. (2012); Chambrone et al. (2011); Chambrone et al. (2013); Chambrone et al. (2011); Fagundes et al. (2019); da Silva et al. (2017); Gomes-Filho et al. (2020); Gomes et al. (2013); Esteves Lima et al. (2021); Artese et al. (2015); Martorano-Fernandes et al. (2020); Peña et al. (2021); da Silva et al. (2021); Nepomuceno et al. (2017); Baeza et al. (2020); Calderaro et al. (2016); Souza et al. (2019); Silveira et al. (2020); Alvarenga et al. (2019); Araújo et al. (2020); Hermont et al. (2014); Souto-Souza et al. (2020); Drumond et al. (2021); Del Rei Daltro Rosa et al. (2021); Porto et al. (2021); Gusman et al. (2018) |
| 42 | China | Yang et al. (2018); Liu T et al. (2021); Gobin et al. (2020); Wei et al. (2021); Guo et al. (2021); Zhu et al. (2017); Chen et al. (2015); Yue et al. (2020); Wang et al. (2020); Cao et al. (2019); Wang et al. (2016); Wu et al. (2020); Zhang et al. (2020); Hua et al. (2016); Peng et al. (2019); Li et al. (2021); Zheng et al. (2021); Tang et al. (2017); Deng et al. (2013); Wang et al. (2021); Chen et al. (2021); Wu et al. (2020); Lü et al. (2011); Liu et al. (2017); Wang et al. (2014); Hu et al. (2021); Wu et al. (2020); Qiao et al. (2020); Xiao et al. (2020); Yang et al. (2018); Zhang Y et al. (2021); Zhang J et al. (2021); Chen et al. (2020); Zhou et al. (2019); Lv et al. (2020); Xu et al. (2020); Ren et al. (2016); Wang et al. (2020); Jiang et al. (2021); Fang et al. (2018); Ji et al. (2021); Sun et al. (2021) |
| 18 | United Kingdom | Muñoz Aguilera et al. (2020); Nadim et al. (2020); Jordão et al. (2020); Simpson et al. (2015); Iheozor-Ejiofor et al. (2017); Joshi et al. (2019); Conde-Agudelo et al. (2008); Simpson et al. (2010); Suvan et al. (2011); Hussain et al. (2020); Nibali et al. (2013); Fuggle et al. (2016); Ratz et al. (2015); Larvin et al. (2021); Orlandi et al. (2014); Rutter-Locher et al. (2017); Hussain et al. (2021); Orlandi et al. (2021) |
| 17 | USA | Ioannidou et al. (2006); Qi et al. (2021); Monje et al. (2017); Abariga et al. (2016); Engebretson et al. (2013); Wijarnpreecha et al. (2020); Demmer et al. (2013); Ungprasert et al. (2017); Schwartz et al. (2018); Bahekar et al. (2007); Hsu et al. (2019); Chaffee et al. (2010); Aminoshariae et al. (2020); Aminoshariae et al. (2018); Lockhart et al. (2019); Oh et al. (2018); Marzouk et al. (2021) |
| 14 | Spain | Galletti et al. (2019); Otero Rey et al. (2019); Tomás et al. (2012); Leira et al. (2017); Leira et al. (2017); Cabanillas-Balsera et al. (2019); Figuero et al. (2013); Lorenzo-Pouso et al. (2021); Romandini et al. (2021); Segura-Egea et al. (2016); Lorenzo-Pouso et al. (2020); Roca-Millan et al. (2018); Manrique-Corredor et al. (2019); Moliner-Sánchez et al. (2020); Bensi et al. (2020); Dioguardi et al. (2019); Corbella et al. (2013); Rapone et al. (2020); Corbella et al. (2016); Corbella et al. (2018); Maisonneuve et al. (2017); Corbella et al. (2012); Dicembrini et al. (2020); Lapo et al. (2021); |
| 10 | Italy | Bensi et al. (2020); Dioguardi et al. (2019); Corbella et al. (2013); Rapone et al. (2020); Corbella et al. (2016); Corbella et al. (2018); Maisonneuve et al. (2017); Corbella et al. (2012); Dicembrini et al. (2020); Lapo et al. (2021) |
| 8 | Australia | Kisely et al. (2015); Kisely et al. (2015); Kapellas et al. (2019); Ali et al. (2020); Kaur et al. (2014); Garde et al. (2019); Jensen et al. (2021); Le et al. (2021) |
| 8 | The Netherlands | Beukers et al. (2021); Ziukaite et al. (2018); Teeuw et al. (2014); Paraskevas et al. (2008); Kunnen et al. (2010); Georgiou et al. (2019); Teeuw et al. (2010); Maarse et al. (2019) |
| 7 | Portugal | Coelho et al. (2020); Machado et al. (2020); Botelho et al. (2018); Machado et al. (2020); Machado et al. (2021); Botelho et al. (2021); Silva et al. (2021) |
| 5 | France | Darnaud et al. (2021); Schmitt et al. (2015); Martin-Cabezas et al. (2016); Darré et al. (2008); Blaizot et al. (2009) |
| 4 | Germany | Papageorgiou et al. (2015); Akcalı et al. (2019); Shang et al. (2021); Stöhr et al. (2021) |
| 4 | Hong Kong | Zhou et al. (2017); Dai et al. (2015); Li et al. (2015); Zhao et al. (2018) |
| 4 | India | Mahajan et al. (2021); Jain et al. (2019); Gupta et al. (2020); Easwaran et al. (2021) |
| 3 | Malaysia | Akram et al. (2016); Gopinath et al. (2020); Zainal Abidin et al. (2021) |
| 3 | Saudi Arabia | Al-Jewair et al. (2015); Farook et al. (2021); AlOtaibi et al. (2021) |
| 3 | Switzerland | Papageorgiou et al. (2017); Maldonado et al. (2018); Koletsi et al. (2021) |
| 2 | Canada | Boutin et al. (2013); Bi et al. (2019) |
| 2 | Colombia | Botero et al. (2020); Rios-Osorio et al. (2020) |
| 2 | Iran | Jalili et al. (2020); Mirzaei et al. (2021) |
| 2 | Norway | Wagle et al. (2018); Skeie et al. (2019) |
| 2 | Romania | Didilescu et al. (2020); Didilescu et al. (2021) |
| 1 | Belgium | Nascimento et al. (2018) |
| 1 | Denmark | Teshome et al. (2016) |
| 1 | Ethiopia | Atieh et al. (2014) |
| 1 | Greece | Martens et al. (2017) |
| 1 | Hungary | Kim et al. (2012) |
| 1 | Lebanon | Chrcanovic et al. (2014) |
| 1 | Morocco | Bouziane et al. (2012) |
| 1 | New Zealand | Polyzos et al. (2010) |
| 1 | Sweden | Ozturk et al. (2021) |
| 1 | Turkey | Andrade et al. (2021) |
