## Supplementary material for "An umbrella review of the evidence linking oral health and systemic health: from the prevalence to clinical and circulating markers": Supplementary data 4.docx

**Supplementary Data 4.** AMSTAR 2 results.

| **Author (Year)** | **1** | **2** | **3** | **4** | **5** | **6** | **7** | **8** | **9** | **10** | **11** | **12** | **13** | **14** | **15** | **16** | **Review Quality** |
| --- | --- | --- | --- | --- | --- | --- | --- | --- | --- | --- | --- | --- | --- | --- | --- | --- | --- |
| Bensi et al. (2020) | Y | PY | N | PY | Y | N | N | PY | Y/Y | N | Y/Y | N | N | Y | NA | Y | Critically Low |
| Wagle et al. (2018) | Y | Y | Y | N | Y | N | N | Y | Y/Y | N | Y/Y | N | N | Y | NA | Y | Critically Low |
| Yang et al. (2018) | Y | N | N | PY | Y | Y | N | PY | Y/Y | N | Y/Y | N | N | Y | NA | Y | Critically Low |
| Coelho et al. (2020) | Y | PY | N | N | Y | N | N | N | Y/Y | N | Y/Y | N | N | Y | N | Y | Critically Low |
| de Lima et al. (2020) | Y | PY | N | N | Y | Y | Y | N | Y/Y | N | Y/Y | N | N | N | NA | Y | Critically Low |
| Kisely et al. (2015) | Y | PY | N | PY | Y | Y | N | Y | Y/Y | N | Y/Y | N | N | Y | Y | Y | Critically Low |
| Liu T et al. (2021) | Y | PY | Y | PY | Y | Y | N | Y | Y/Y | N | Y/Y | N | N | Y | Y | Y | Critically Low |
| Skeie et al. (2019) | Y | PY | Y | N | Y | Y | N | Y | Y/Y | N | Y/Y | N | Y | Y | Y | Y | Critically Low |
| Cademartori et al. (2018) | Y | Y | Y | PY | N | Y | Y | PY | Y/Y | N | Y/Y | N | N | Y | NA | Y | Low |
| Zhou et al. (2017) | Y | PY | N | N | Y | Y | N | PY | Y/Y | N | Y/Y | N | N | N | Y | Y | Critically Low |
| Didilescu et al. (2020) | Y | PY | N | N | Y | N | N | PY | Y/Y | N | Y/Y | N | N | N | NA | N | Critically Low |
| Mahajan et al. (2021) | Y | N | Y | N | Y | Y | N | PY | Y/Y | N | Y/Y | N | N | Y | Y | Y | Critically Low |
| Beukers et al. (2021) | Y | Y | Y | N | Y | N | N | Y | Y/Y | N | Y/Y | N | N | Y | Y | Y | Critically Low |
| Papageorgiou et al. (2017) | Y | Y | Y | Y | Y | Y | Y | Y | Y/Y | N | Y/Y | N | Y | Y | NA | Y | Moderate |
| Akcalı et al. (2019) | Y | Y | N | PY | Y | Y | Y | Y | Y/Y | N | Y/Y | N | Y | Y | Y | Y | Moderate |
| Cerutti-Kopplin et al. (2016) | Y | Y | Y | PY | Y | Y | N | Y | Y/Y | N | Y/Y | N | Y | Y | NA | Y | Low |
| Dai et al. (2015) | Y | N | Y | N | Y | Y | Y | Y | Y/Y | N | Y/Y | N | N | Y | NA | Y | Critically Low |
| Nascimento et al. (2018) | Y | Y | Y | PY | Y | Y | Y | Y | Y/Y | N | Y/Y | N | Y | Y | Y | Y | Moderate |
| Li et al. (2015) | Y | N | Y | Y | Y | Y | Y | Y | Y/Y | N | Y/Y | N | N | Y | Y | Y | Critically Low |
| Galletti et al. (2019) | Y | PY | Y | Y | Y | Y | N | Y | Y/Y | N | Y/Y | Y | Y | Y | NA | Y | Low |
| Muñoz Aguilera et al. (2020) | Y | Y | Y | Y | Y | Y | N | Y | Y/Y | N | Y/Y | Y | Y | Y | Y | Y | Low |
| Kisely et al. (2015) | Y | PY | N | PY | Y | N | N | Y | Y/Y | N | Y/Y | N | N | Y | Y | Y | Critically Low |
| Kapellas et al. (2019) | Y | Y | Y | PY | Y | N | N | Y | Y/Y | N | Y/Y | Y | Y | Y | Y | Y | Low |
| Nascimento et al. (2015) | Y | PY | Y | PY | Y | Y | Y | Y | Y/Y | N | Y/Y | N | Y | Y | NA | Y | Moderate |
| Nadim et al. (2020) | Y | N | Y | PY | N | N | N | Y | Y/Y | N | Y/Y | N | N | N | Y | Y | Critically Low |
| Ioannidou et al. (2006) | Y | N | Y | N | Y | N | N | Y | Y/Y | N | Y/Y | Y | Y | Y | N | N | Critically Low |
| Jerônimo et al. (2020) | Y | PY | Y | PY | Y | N | N | Y | Y/Y | N | Y/Y | N | N | Y | N | N | Critically Low |
| Darnaud et al. (2021) | Y | N | Y | Y | Y | N | Y | Y | Y/Y | N | Y/Y | N | N | Y | N | Y | Critically Low |
| Didilescu et al. (2021) | Y | PY | Y | PY | N | N | N | Y | Y/Y | N | Y/Y | N | N | Y | NA | Y | Critically Low |
| Gobin et al. (2020) | Y | Y | Y | N | Y | Y | N | Y | Y/Y | Y | Y/Y | N | N | Y | Y | Y | Critically Low |
| Wei et al. (2021) | Y | PY | Y | PY | Y | Y | N | Y | Y/Y | N | Y/Y | Y | Y | Y | NA | Y | Low |
| Guo et al. (2021) | Y | PY | Y | PY | N | Y | N | Y | Y/Y | N | Y/Y | N | N | Y | NA | Y | Critically Low |
| Akram et al. (2016) | Y | PY | Y | PY | Y | Y | Y | Y | Y/Y | N | Y/Y | N | N | Y | N | Y | Critically Low |
| Zhu et al. (2017) | Y | Y | Y | PY | Y | Y | N | N | Y/Y | N | Y/Y | N | Y | Y | Y | Y | Low |
| Qi et al. (2021) | Y | PY | Y | N | N | Y | N | Y | Y/Y | N | Y/Y | Y | Y | Y | Y | Y | Low |
| Jordão et al. (2020) | Y | N | Y | N | Y | N | N | Y | Y/Y | N | Y/Y | N | N | Y | Y | Y | Critically Low |
| Teshome et al. (2016) | Y | PY | Y | PY | Y | Y | N | Y | Y/Y | N | Y/Y | N | Y | Y | N | Y | Low |
| Simpson et al. (2015) | Y | Y | Y | Y | Y | Y | Y | Y | Y/Y | Y | Y/Y | Y | Y | Y | Y | Y | High |
| Iheozor-Ejiofor et al. (2017) | Y | PY | Y | PY | Y | Y | Y | Y | Y/Y | Y | Y/Y | Y | Y | Y | Y | Y | High |
| Galdino et al. (2021) | Y | Y | Y | PY | Y | Y | Y | Y | Y/Y | N | Y/Y | N | N | Y | Y | Y | Low |
| Chen et al. (2015) | Y | Y | Y | PY | Y | N | N | Y | Y/Y | N | Y/Y | N | N | Y | Y | Y | Critically Low |
| Ali et al. (2020) | Y | Y | Y | PY | Y | N | N | Y | Y/Y | N | Y/Y | Y | Y | Y | NA | Y | Low |
| Nascimento et al. (2016) | Y | PY | Y | N | Y | Y | Y | Y | Y/Y | N | Y/Y | N | Y | Y | N | Y | Critically Low |
| Yue et al. (2020) | Y | PY | Y | N | Y | Y | N | PY | Y/Y | N | Y/Y | N | Y | Y | Y | Y | Critically Low |
| Wang et al. (2020) | Y | PY | Y | N | Y | Y | Y | Y | Y/Y | N | Y/Y | Y | Y | Y | Y | Y | Low |
| Otero Rey et al. (2019) | Y | PY | Y | N | Y | Y | N | Y | Y/Y | N | Y/Y | N | N | Y | N | Y | Critically Low |
| Dioguardi et al. (2019) | Y | N | Y | N | Y | Y | N | Y | Y/Y | N | Y/Y | N | N | Y | NA | Y | Critically Low |
| Cao et al. (2019) | Y | Y | Y | PY | Y | Y | Y | Y | Y/Y | N | Y/Y | N | N | Y | Y | Y | Low |
| Corbella et al. (2013) | Y | N | Y | PY | N | N | N | Y | Y/Y | N | Y/Y | N | N | Y | N | Y | Critically Low |
| Wang et al. (2016) | Y | Y | Y | Y | Y | Y | Y | Y | Y/Y | Y | Y/Y | Y | Y | Y | NA | Y | High |
| Gopinath et al. (2020) | Y | Y | Y | N | Y | Y | N | Y | Y/Y | N | Y/Y | N | N | N | Y | Y | Critically Low |
| Rapone et al. (2020) | Y | N | Y | N | Y | Y | N | Y | N/N | N | Y/Y | N | Y | N | N | Y | Critically Low |
| Wu et al. (2020) | Y | Y | Y | PY | N | Y | N | Y | Y/Y | N | Y/Y | N | N | Y | Y | Y | Critically Low |
| Botero et al. (2020) | Y | Y | N | N | Y | Y | Y | Y | Y/Y | N | Y/Y | Y | Y | Y | Y | Y | Low |
| Zhang et al. (2020) | Y | PY | Y | PY | Y | Y | N | N | Y/Y | N | Y/Y | N | N | Y | N | Y | Critically Low |
| Schmitt et al. (2015) | Y | PY | Y | N | Y | N | Y | Y | Y/Y | N | Y/Y | Y | Y | Y | NA | Y | Low |
| Atieh et al. (2014) | Y | PY | Y | N | N | N | N | Y | Y/Y | N | Y/Y | N | N | Y | N | Y | Critically Low |
| Corbella et al. (2016) | Y | N | Y | PY | Y | Y | Y | Y | Y/Y | N | Y/Y | Y | Y | Y | N | N | Critically Low |
| Hua et al. (2016) | Y | Y | Y | PY | Y | Y | Y | Y | Y/Y | Y | Y/Y | Y | Y | Y | Y | Y | High |
| Araújo et al. (2016) | Y | PY | Y | N | Y | Y | N | Y | Y/Y | N | Y/Y | N | N | Y | N | Y | Critically Low |
| Ziukaite et al. (2018) | Y | Y | Y | N | Y | Y | N | Y | Y/Y | N | Y/Y | N | N | Y | Y | Y | Critically Low |
| Ferreira et al. (2019) | Y | Y | Y | Y | N | N | Y | Y | Y/Y | N | Y/Y | N | N | N | NA | Y | Low |
| Teeuw et al. (2014) | Y | Y | Y | PY | Y | Y | Y | Y | Y/Y | N | Y/Y | N | N | Y | NA | Y | Low |
| Papageorgiou et al. (2015) | Y | Y | Y | Y | Y | Y | Y | Y | Y/Y | N | Y/Y | N | Y | Y | NA | Y | Moderate |
| Tomás et al. (2012) | Y | PY | Y | PY | Y | N | N | Y | Y/Y | N | Y/Y | N | N | Y | NA | Y | Critically Low |
| Peng et al. (2019) | Y | PY | Y | PY | Y | Y | Y | Y | Y/Y | N | Y/Y | Y | N | Y | Y | Y | Low |
| Ferreira et al. (2019) | Y | Y | Y | Y | Y | Y | Y | Y | Y/Y | N | Y/Y | Y | Y | Y | N | Y | Low |
| da Silva et al. (2021) | Y | Y | Y | N | Y | Y | N | Y | Y/Y | N | Y/Y | N | N | Y | N | Y | Critically Low |
| Daudt et al. (2018) | Y | Y | Y | PY | Y | Y | N | Y | Y/Y | N | Y/Y | N | N | Y | Y | Y | Critically Low |
| Zhao et al. (2018) | Y | N | Y | N | Y | N | N | PY | Y/Y | N | Y/Y | N | Y | Y | Y | Y | Critically Low |
| Moraschini et al. (2016) | Y | PY | Y | Y | Y | N | N | Y | Y/Y | N | Y/Y | N | Y | N | Y | Y | Low |
| Leira et al. (2017) | Y | Y | Y | PY | Y | Y | N | Y | Y/Y | N | Y/Y | Y | Y | Y | NA | Y | Low |
| Moraschini et al. (2018) | Y | PY | Y | Y | Y | Y | N | Y | Y/Y | N | Y/Y | N | Y | Y | N | Y | Critically Low |
| Joshi et al. (2019) | Y | PY | N | N | Y | Y | N | Y | Y/Y | N | Y/Y | N | N | Y | N | N | Critically Low |
| Conde-Agudelo et al. (2008) | Y | PY | Y | PY | N | N | N | Y | N/N | N | Y/Y | Y | Y | Y | Y | Y | Critically Low |
| Simpson et al. (2010) | Y | Y | Y | Y | Y | Y | Y | Y | Y/Y | Y | Y/Y | Y | Y | Y | Y | Y | High |
| Boutin et al. (2013) | Y | PY | Y | PY | Y | Y | N | Y | Y/Y | N | Y/Y | Y | Y | Y | N | Y | Critically Low |
| Suvan et al. (2011) | Y | Y | Y | Y | Y | Y | N | Y | Y/Y | N | Y/Y | N | N | N | Y | Y | Critically Low |
| Moura-Grec et al. (2014) | Y | N | Y | N | Y | Y | N | Y | N/N | N | Y/Y | N | Y | Y | N | N | Critically Low |
| Machado et al. (2020) | Y | Y | Y | Y | Y | Y | N | Y | Y/Y | N | Y/Y | Y | Y | Y | NA | Y | Low |
| de Oliveira Ferreira et al. (2019) | Y | N | Y | Y | Y | N | Y | Y | Y/Y | N | Y/Y | N | N | Y | NA | Y | Critically Low |
| Monje et al. (2017) | Y | PY | Y | Y | Y | Y | Y | Y | Y/Y | N | Y/Y | Y | Y | Y | NA | Y | High |
| Martens et al. (2017) | Y | Y | Y | Y | Y | Y | N | Y | Y/Y | N | Y/Y | N | N | Y | Y | Y | Critically Low |
| Esteves Lima et al. (2016) | Y | Y | Y | PY | Y | Y | Y | Y | Y/Y | N | Y/Y | N | Y | Y | NA | Y | Critically Low |
| Li et al. (2021) | Y | PY | Y | PY | N | N | N | Y | Y/Y | N | Y/Y | N | N | Y | Y | Y | Critically Low |
| Rosa et al. (2012) | Y | N | Y | PY | Y | Y | N | Y | Y/Y | N | Y/Y | N | N | Y | Y | N | Critically Low |
| Leira et al. (2017) | Y | Y | Y | PY | Y | Y | N | Y | Y/Y | N | Y/Y | N | N | Y | NA | Y | Critically Low |
| Hussain et al. (2020) | Y | Y | Y | Y | Y | Y | Y | Y | Y/Y | N | Y/Y | Y | Y | Y | NA | Y | High |
| Botelho et al. (2018) | Y | PY | Y | N | Y | Y | N | Y | Y/Y | N | Y/Y | N | Y | Y | Y | Y | Critically Low |
| Zheng et al. (2021) | Y | Y | Y | N | Y | N | N | PY | Y/Y | N | Y/Y | Y | Y | Y | Y | Y | Critically Low |
| Chambrone et al. (2011) | Y | Y | Y | Y | Y | Y | Y | Y | Y/Y | Y | Y/Y | N | Y | Y | NA | Y | High |
| Jain A et al. (2019) | Y | N | Y | N | Y | Y | N | Y | Y/Y | N | Y/Y | N | Y | N | Y | Y | Critically Low |
| Martin-Cabezas et al. (2016) | Y | N | Y | N | N | N | N | Y | Y/Y | N | Y/Y | N | N | Y | N | Y | Critically Low |
| Shang et al. (2021) | Y | Y | Y | PY | Y | N | N | Y | N/N | N | Y/Y | Y | Y | Y | Y | N | Critically Low |
| Chambrone et al. (2013) | Y | Y | Y | PY | Y | Y | Y | Y | Y/Y | N | Y/Y | Y | Y | Y | NA | Y | High |
| Chambrone et al. (2011) | Y | PY | Y | Y | Y | Y | Y | Y | Y/Y | Y | Y/Y | N | Y | Y | NA | Y | High |
| Paraskevas et al. (2008) | Y | N | Y | N | Y | N | Y | Y | Y/Y | N | Y/Y | N | N | N | N | Y | Critically Low |
| Fagundes et al. (2019) | Y | PY | Y | PY | Y | Y | N | Y | Y/Y | N | Y/Y | N | N | Y | NA | Y | Critically Low |
| da Silva et al. (2017) | Y | PY | Y | PY | Y | Y | Y | Y | Y/Y | N | Y/Y | N | N | Y | N | Y | Critically Low |
| Bi et al. (2019) | Y | N | Y | PY | Y | Y | N | Y | Y/Y | N | Y/Y | N | N | Y | Y | Y | Critically Low |
| Cabanillas-Balsera et al. (2019) | Y | PY | Y | PY | Y | N | N | N | Y/Y | N | Y/Y | N | N | N | N | Y | Critically Low |
| Nibali et al. (2013) | Y | Y | Y | PY | Y | Y | N | Y | Y/Y | N | Y/Y | Y | N | Y | Y | Y | Critically Low |
| Gomes-Filho et al. (2020) | Y | PY | Y | PY | Y | Y | N | Y | Y/Y | N | Y/Y | N | N | Y | Y | Y | Critically Low |
| Kim et al. (2012) | Y | PY | Y | PY | Y | Y | N | Y | Y/Y | N | Y/Y | N | N | Y | Y | Y | Low |
| Maldonado et al. (2018) | Y | N | Y | N | Y | N | N | Y | Y/Y | N | Y/Y | N | N | Y | N | Y | Critically Low |
| Corbella et al. (2018) | Y | Y | Y | PY | Y | Y | Y | Y | Y/Y | N | Y/Y | Y | Y | Y | N | Y | Low |
| Chrcanovic et al. (2014) | Y | Y | Y | PY | Y | Y | N | Y | Y/Y | N | Y/Y | N | N | Y | Y | Y | Critically Low |
| Tang et al. (2017) | Y | PY | Y | PY | Y | Y | N | Y | Y/Y | Y | Y/Y | N | N | Y | Y | Y | Critically Low |
| Deng et al. (2013) | Y | N | Y | PY | Y | Y | N | Y | Y/Y | N | Y/Y | N | N | Y | N | N | Critically Low |
| Gupta et al. (2020) | Y | PY | Y | N | Y | Y | Y | Y | Y/Y | N | Y/Y | N | N | Y | Y | Y | Critically Low |
| Kaur et al. (2014) | Y | N | Y | PY | Y | N | N | Y | Y/Y | N | Y/Y | N | N | N | N | Y | Critically Low |
| Gomes et al. (2013) | Y | PY | Y | PY | Y | N | N | Y | Y/Y | N | Y/Y | N | N | Y | N | Y | Critically Low |
| Wang et al. (2021) | Y | PY | Y | PY | Y | Y | N | Y | Y/Y | N | Y/Y | Y | Y | Y | Y | Y | Low |
| Esteves Lima et al. (2021) | Y | PY | Y | PY | Y | N | Y | Y | Y/Y | N | Y/Y | N | N | N | N | Y | Critically Low |
| Abariga et al. (2016) | Y | N | Y | N | N | Y | N | Y | Y/Y | N | Y/Y | Y | Y | Y | Y | Y | Critically Low |
| Engebretson et al. (2013) | Y | PY | Y | N | N | N | N | Y | Y/Y | N | Y/Y | N | N | Y | Y | Y | Critically Low |
| Figuero et al. (2013) | Y | PY | Y | N | Y | Y | N | Y | Y/Y | Y | Y/Y | N | N | Y | Y | Y | Critically Low |
| Kunnen et al. (2010) | Y | N | Y | N | Y | N | Y | Y | Y/Y | N | Y/Y | Y | Y | Y | N | Y | Critically Low |
| Machado et al. (2020) | Y | PY | Y | Y | Y | Y | N | Y | Y/Y | N | Y/Y | Y | Y | Y | NA | Y | High |
| Georgiou et al. (2019) | Y | PY | Y | PY | Y | N | Y | Y | Y/Y | Y | Y/Y | N | N | Y | N | Y | Critically Low |
| Garde et al. (2019) | Y | Y | Y | PY | Y | N | N | Y | Y/Y | N | Y/Y | N | N | Y | N | Y | Critically Low |
| Lorenzo-Pouso et al. (2021) | Y | Y | Y | PY | Y | Y | N | Y | Y/Y | N | Y/Y | Y | Y | Y | Y | Y | Low |
| Romandini et al. (2021) | Y | Y | Y | PY | Y | Y | Y | Y | Y/Y | N | Y/Y | N | N | Y | Y | Y | Low |
| Artese et al. (2015) | Y | Y | Y | N | Y | Y | N | Y | Y/Y | N | Y/Y | N | N | Y | NA | Y | Critically Low |
| Segura-Egea et al. (2016) | Y | Y | Y | PY | Y | N | Y | Y | Y/Y | N | Y/Y | N | N | Y | N | Y | Critically Low |
| Chen et al. (2021) | Y | PY | Y | PY | Y | Y | N | Y | Y/Y | N | Y/Y | Y | Y | Y | Y | Y | Low |
| Maisonneuve et al. (2017) | Y | N | Y | PY | Y | Y | N | Y | N/N | N | Y/Y | N | N | Y | Y | Y | Critically Low |
| Bouziane et al. (2012) | Y | PY | Y | Y | Y | Y | N | Y | Y/Y | N | Y/Y | N | N | Y | NA | Y | Critically Low |
| Darré et al. (2008) | Y | N | Y | Y | Y | N | N | Y | Y/Y | N | Y/Y | N | N | Y | Y | Y | Critically Low |
| Machado et al. (2021) | Y | Y | Y | Y | Y | Y | Y | Y | Y/Y | Y | Y/Y | Y | Y | Y | Y | Y | High |
| Wu et al. (2020) | Y | Y | Y | PY | N | N | N | Y | Y/Y | N | Y/Y | N | N | Y | N | Y | Critically Low |
| Lü et al. (2011) | Y | N | Y | N | Y | N | N | Y | Y/Y | N | Y/Y | N | Y | Y | N | N | Critically Low |
| Corbella et al. (2012) | Y | N | Y | N | Y | N | N | Y | N/N | N | Y/Y | N | N | N | Y | N | Critically Low |
| Wijarnpreecha et al. (2020) | Y | PY | Y | PY | Y | Y | N | Y | Y/Y | N | Y/Y | N | N | Y | Y | Y | Critically Low |
| Lorenzo-Pouso et al. (2020) | Y | PY | Y | N | Y | Y | N | Y | Y/Y | N | Y/Y | N | Y | Y | N | Y | Critically Low |
| Demmer et al. (2013) | Y | PY | Y | Y | Y | Y | N | Y | Y/Y | N | Y/Y | N | N | Y | Y | Y | Critically Low |
| Polyzos et al. (2010) | Y | PY | Y | N | N | Y | N | Y | Y/Y | N | Y/Y | N | Y | N | Y | Y | Critically Low |
| Martorano-Fernandes et al. (2020) | Y | PY | Y | Y | N | Y | N | Y | Y/Y | N | Y/Y | Y | Y | Y | N | Y | Critically Low |
| Peña et al. (2021) | Y | PY | Y | N | N | N | Y | Y | Y/Y | N | Y/Y | N | N | N | Y | Y | Critically Low |
| Botelho et al. (2021) | Y | Y | Y | Y | Y | Y | Y | Y | Y/Y | Y | Y/Y | Y | Y | Y | Y | Y | High |
| Roca-Millan et al. (2018) | Y | N | N | N | N | N | N | Y | Y/Y | N | Y/Y | N | N | Y | NA | Y | Critically Low |
| Jensen et al. (2021) | Y | PY | N | N | Y | N | N | Y | Y/Y | N | Y/Y | N | Y | N | Y | Y | Critically Low |
| Manrique-Corredor et al. (2019) | Y | N | Y | N | Y | Y | N | Y | Y/Y | N | Y/Y | N | Y | Y | Y | Y | Critically Low |
| Ungprasert et al. (2017) | Y | PY | Y | N | Y | Y | N | Y | Y/Y | N | Y/Y | N | N | Y | Y | Y | Critically Low |
| Liu et al. (2017) | Y | Y | Y | PY | Y | N | N | Y | N/N | N | Y/Y | Y | Y | N | N | Y | Critically Low |
| Teeuw et al. (2010) | Y | PY | Y | N | N | N | Y | Y | Y/Y | N | Y/Y | N | N | N | NA | Y | Critically Low |
| Farook et al. (2021) | Y | Y | Y | PY | Y | Y | N | Y | Y/Y | N | Y/Y | N | Y | Y | NA | Y | Low |
| Wang et al. (2014) | Y | N | Y | PY | Y | Y | N | Y | Y/Y | N | Y/Y | N | N | Y | Y | Y | Critically Low |
| Hu et al. (2021) | Y | Y | Y | Y | Y | Y | N | Y | Y/Y | N | Y/Y | N | N | Y | NA | Y | Critically Low |
| Schwartz et al. (2018) | Y | PY | Y | PY | Y | Y | N | Y | Y/Y | N | Y/Y | Y | Y | Y | N | Y | Critically Low |
| Bahekar et al. (2007) | Y | N | Y | PY | Y | Y | N | Y | Y/Y | N | Y/Y | Y | Y | N | N | N | Critically Low |
| Hsu et al. (2019) | Y | Y | Y | PY | Y | Y | Y | Y | Y/Y | N | Y/Y | N | N | Y | Y | Y | Low |
| Fuggle et al. (2016) | Y | PY | Y | N | Y | Y | N | Y | Y/Y | N | Y/Y | N | N | Y | N | Y | Critically Low |
| Stöhr et al. (2021) | Y | Y | Y | PY | Y | Y | Y | Y | Y/Y | N | Y/Y | N | N | Y | Y | Y | Low |
| Ratz et al. (2015) | Y | PY | N | PY | Y | N | N | Y | Y/Y | Y | Y/Y | N | N | Y | NA | Y | Critically Low |
| Wu et al. (2020) | Y | N | Y | PY | N | N | Y | Y | Y/Y | N | Y/Y | N | N | Y | Y | Y | Critically Low |
| Al-Jewair et al. (2015) | Y | Y | Y | Y | Y | Y | N | Y | Y/Y | N | Y/Y | N | N | N | NA | Y | Critically Low |
| Chaffee et al. (2010) | Y | PY | Y | PY | Y | Y | N | Y | Y/Y | N | Y/Y | Y | Y | Y | Y | Y | Low |
| da Silva et al. (2021) | Y | PY | Y | PY | Y | N | Y | Y | Y/Y | N | Y/Y | N | N | N | Y | Y | Low |
| Larvin et al. (2021) | Y | PY | Y | N | N | N | N | Y | Y/Y | N | Y/Y | N | N | Y | Y | Y | Critically Low |
| Nepomuceno et al. (2017) | Y | PY | Y | N | N | Y | N | Y | Y/Y | N | Y/Y | N | Y | Y | Y | Y | Critically Low |
| Qiao et al. (2020) | Y | N | Y | PY | Y | Y | N | Y | Y/Y | N | Y/Y | N | N | Y | Y | Y | Critically Low |
| Dicembrini et al. (2020) | Y | PY | Y | N | Y | Y | N | Y | Y/Y | N | Y/Y | N | N | N | Y | Y | Critically Low |
| Xiao et al. (2020) | Y | Y | N | PY | Y | Y | N | Y | Y/Y | N | Y/Y | N | N | Y | Y | Y | Critically Low |
| Yang et al. (2018) | Y | PY | N | N | Y | Y | N | Y | Y/Y | N | Y/Y | N | N | N | N | Y | Critically Low |
| Orlandi et al. (2014) | Y | Y | Y | Y | Y | Y | N | Y | Y/Y | N | Y/Y | Y | Y | Y | Y | Y | Low |
| Zhang Y et al. (2021) | Y | PY | Y | N | N | Y | Y | Y | Y/Y | N | Y/Y | N | N | Y | N | Y | Critically Low |
| Zhang J et al. (2021) | Y | Y | Y | PY | Y | Y | N | Y | Y/Y | N | Y/Y | N | N | Y | N | Y | Critically Low |
| Moliner-Sánchez et al. (2020) | Y | PY | Y | PY | Y | Y | N | Y | Y/Y | N | Y/Y | N | N | Y | Y | Y | Critically Low |
| Baeza et al. (2020) | Y | Y | Y | N | Y | Y | Y | Y | Y/Y | N | Y/Y | N | N | Y | Y | Y | Critically Low |
| Rutter-Locher et al. (2017) | Y | PY | Y | N | Y | Y | N | Y | Y/Y | N | Y/Y | N | N | Y | N | Y | Critically Low |
| Chen et al. (2020) | Y | PY | Y | PY | N | Y | N | Y | Y/Y | N | Y/Y | Y | Y | Y | Y | Y | Low |
| Koletsi et al. (2021) | Y | PY | Y | Y | Y | Y | N | Y | Y/Y | N | Y/Y | Y | Y | Y | Y | N | Moderate |
| Ozturk et al. (2021) | Y | Y | Y | PY | N | N | N | Y | Y/Y | N | Y/Y | N | N | Y | N | Y | Critically Low |
| Zhou et al. (2019) | Y | Y | Y | N | N | Y | N | Y | Y/Y | N | Y/Y | N | N | Y | N | Y | Critically Low |
| Lv et al. (2020) | Y | PY | Y | PY | Y | Y | N | Y | Y/Y | N | Y/Y | N | N | Y | Y | Y | Critically Low |
| Xu et al. (2020) | Y | PY | Y | PY | Y | Y | N | Y | Y/Y | N | Y/Y | Y | Y | Y | N | Y | Critically Low |
| Silva et al. (2021) | Y | PY | Y | PY | Y | Y | N | Y | Y/Y | N | Y/Y | N | Y | N | N | Y | Critically Low |
| Calderaro et al. (2016) | Y | N | N | PY | Y | Y | N | N | Y/Y | N | Y/Y | N | N | Y | N | Y | Critically Low |
| Ren et al. (2016) | Y | Y | Y | Y | Y | Y | Y | Y | Y/Y | N | Y/Y | Y | N | Y | NA | N | Moderate |
| Zainal Abidin et al. (2021) | Y | Y | N | PY | N | N | N | Y | Y/Y | N | Y/Y | N | N | Y | N | Y | Critically Low |
| Hussain et al. (2021) | Y | Y | Y | Y | Y | Y | N | Y | Y/Y | N | Y/Y | Y | N | N | NA | Y | Critically Low |
| Mirzaei et al. (2021) | Y | Y | Y | PY | Y | N | N | Y | Y/Y | N | Y/Y | Y | Y | Y | Y | Y | Low |
| Maarse et al. (2019) | Y | Y | Y | N | Y | N | N | Y | Y/Y | N | Y/Y | N | N | N | N | N | Critically Low |
| Souza et al. (2019) | Y | Y | Y | PY | Y | Y | Y | Y | Y/Y | N | Y/Y | N | N | Y | N | Y | Critically Low |
| Jalili et al. (2020) | Y | N | N | N | Y | Y | N | N | Y/Y | N | Y/Y | N | N | Y | N | Y | Critically Low |
| Silveira et al. (2020) | Y | PY | N | Y | Y | Y | N | Y | Y/Y | N | Y/Y | N | Y | Y | Y | Y | Low |
| Aminoshariae et al. (2020) | Y | PY | N | N | N | N | Y | Y | Y/Y | N | Y/Y | Y | N | Y | NA | Y | Critically Low |
| Aminoshariae et al. (2018) | Y | PY | N | N | N | N | Y | Y | Y/Y | N | Y/Y | Y | N | Y | NA | Y | Critically Low |
| Alvarenga et al. (2019) | Y | Y | Y | Y | Y | Y | Y | Y | Y/Y | N | Y/Y | Y | Y | Y | Y | Y | High |
| Araújo et al. (2020) | Y | Y | Y | PY | Y | Y | N | N | Y/Y | N | Y/Y | N | N | Y | Y | Y | Critically Low |
| Hermont et al. (2014) | Y | N | Y | Y | Y | Y | Y | Y | Y/Y | N | Y/Y | N | N | Y | NA | Y | Critically Low |
| Lockhart et al. (2019) | Y | Y | Y | PY | Y | Y | Y | Y | Y/Y | N | Y/Y | Y | Y | Y | N | Y | Low |
| Wang et al. (2020) | Y | N | Y | N | N | N | N | Y | N/N | N | Y/Y | N | N | Y | Y | Y | Critically Low |
| Jiang et al. (2021) | Y | Y | N | Y | N | N | N | Y | Y/Y | N | Y/Y | N | N | Y | Y | Y | Critically Low |
| Fang et al. (2018) | Y | PY | N | N | N | N | N | Y | Y/Y | N | Y/Y | N | N | Y | Y | Y | Critically Low |
| Souto-Souza et al. (2020) | Y | PY | Y | PY | Y | N | N | PY | Y/Y | N | Y/Y | N | Y | Y | Y | Y | Low |
| Rios-Osorio et al. (2020) | Y | N | Y | Y | N | Y | N | Y | Y/Y | N | Y/Y | Y | N | Y | NA | Y | Critically Low |
| Blaizot et al. (2009) | Y | N | Y | PY | Y | Y | N | Y | Y/Y | N | Y/Y | N | N | Y | Y | N | Critically Low |
| Oh et al. (2018) | Y | PY | N | N | Y | Y | N | Y | Y/Y | N | Y/Y | N | N | Y | Y | Y | Critically Low |
| AlOtaibi et al. (2021) | Y | N | Y | N | N | N | N | Y | N/N | N | Y/Y | N | N | N | NA | Y | Critically Low |
| Easwaran et al. (2021) | Y | Y | Y | Y | Y | Y | Y | Y | Y/Y | N | Y/Y | N | N | N | NA | Y | Low |
| Ji et al. (2021) | Y | N | Y | N | N | N | N | Y | N/N | N | Y/Y | N | N | Y | NA | Y | Critically Low |
| Sun et al. (2021) | Y | PY | Y | PY | Y | Y | N | Y | Y/Y | N | Y/Y | N | N | Y | NA | Y | Critically Low |
| Drumond et al. (2021) | Y | PY | Y | PY | Y | Y | Y | Y | Y/Y | N | Y/Y | N | N | Y | NA | Y | Low |
| Le et al. (2021) | Y | PY | Y | PY | Y | N | N | Y | Y/Y | N | Y/Y | N | N | N | Y | Y | Critically Low |
| Marzouk et al. (2021) | Y | PY | N | Y | Y | Y | Y | N | Y/Y | N | Y/Y | N | Y | Y | NA | Y | Moderate |
| Del Rei Daltro Rosa et al. (2021) | Y | PY | Y | PY | Y | N | Y | Y | Y/Y | N | Y/Y | N | N | Y | NA | Y | Low |
| Porto et al. (2021) | Y | PY | Y | N | Y | Y | N | Y | Y/Y | Y | Y/Y | N | Y | N | N | N | Critically Low |
| Orlandi et al. (2021) | Y | Y | Y | PY | Y | Y | Y | Y | Y/Y | Y | Y/Y | Y | Y | Y | Y | Y | High |
| Lapo et al. (2021) | Y | PY | Y | PY | Y | Y | N | Y | Y/Y | N | Y/Y | N | N | Y | NA | Y | Critically Low |
| Andrade et al. (2021) | Y | PY | Y | Y | Y | Y | N | Y | Y/Y | N | Y/Y | N | N | Y | NA | Y | Critically Low |
| Gusman et al. (2018) | Y | PY | Y | N | Y | Y | Y | Y | Y/Y | N | Y/Y | N | N | Y | NA | Y | Critically Low |

N—No, Y—Yes, PY—Partial Yes. 1. Research questions and inclusion criteria? 2. Review methods established a priori? 3. Explanation of their selection literature search strategy? 4. Did the review authors use a comprehensive literature search strategy? 5. Study selection performed in duplicate? 6. Data selection performed in duplicate? 7. List of excluded studies and exclusions justified? 8. Description of the included studies in adequate detail? 9. Satisfactory technique for assessing the risk of bias (RoB)? 10. Report on the sources of funding for the studies included in the review? 11. If meta-analysis was performed, did the review authors use appropriate methods for statistical combination of results? 12. If meta-analysis was performed, did the review authors assess the potential impact of RoB? 13. RoB accounted when interpreting/discussing the results of the review? 14. Did the review authors provide a satisfactory explanation for, and discussion of, any heterogeneity observed in the results of the review? 15. If they performed quantitative synthesis, was publication bias performed? 16. Did the review authors report any potential sources of conflict of interest, including funding sources?.
